# Novel Cognitive Behavioral Therapy App for Depression in Mild Traumatic Brain Injury: A Randomized Controlled Trial

**DOI:** 10.64898/2026.09.24.26363933

**Authors:** J. Bronte Emery, Leonard Matheson, Olivia M. Shaw, Noushin Mannan, Martin R. Cota, Adele Fu, Thaddeus J. Haight, David L. Brody

**Affiliations:** The Henry M. Jackson Foundation for the Advancement of Military Medicine, Inc., Bethesda, MD, United States of America; Military Traumatic Brain Injury Initiative, formerly known as the Center for Neuroscience and Regenerative Medicine, Bethesda, MD, United States of America; EPIC Neurorehabilitation & Psychology Services, Inc., Chico, CA, United States of America; Department of Neurology, Uniformed Services University of the Health Sciences, Bethesda, MD, United States of America

**Author notes:** **Disclaimer**: The views, information or content, and conclusions presented do not necessarily represent the official position or policy of, nor should any official endorsement be inferred on the part of, the Uniformed Services University, the Department of Defense, Department of War, the U.S. Government or the Henry M. Jackson Foundation for the Advancement of Military Medicine, Inc. Email addresses.

**Keywords:** Traumatic brain injury, concussion, depression, cognitive behavioral therapy, randomized controlled trial, insomnia

## Abstract

**Background:** Depression symptoms following traumatic brain injury (TBI) are common and strongly associated with adverse overall outcomes in military service members, veterans, and civilians. Cognitive behavioral therapy for depression (CBT-D) is considered a first-line treatment for depression symptoms in many contexts. However, costs associated with CBT-D can be substantial, and there are insufficient numbers of providers trained to perform CBT-D. In contradistinction, digital therapeutic interventions such as smartphone apps have virtually unlimited availability and low per-patient costs. However, evidence for efficacy of digital interventions providing CBT-D in the context of TBI is limited. The aims of this study were to design, develop, and test a novel smartphone app to provide CBT-D intended for military service members with depression symptoms and history of mild TBI.

**Results:** We designed a novel smartphone app featuring digital avatar coaches to provide CBT-D intended for military service members with depression symptoms and history of mild TBI. Key features and goals of the 12 weekly CBT lessons included stigma minimization, personal engagement, and 1-2 hours per week of personal challenge homework assignments. We performed a randomized controlled trial initially involving military service members and veterans. Inclusion criteria were later relaxed to allow for civilian participation due to practical constraints. The control group received an app providing educational lessons about concussions and depression which did not include the therapeutic components of CBT. There was no difference between the groups randomized to CBT-D (n=19) vs. educational app control (n=23) in self-reported depression symptoms. Nevertheless, there was a statistically significant benefit of the CBT-D intervention in self-reported insomnia severity at the 16-week follow-up (four weeks post intervention) assessment (Cohen’s D=0.45, p=0.0064). There were also statistically significant positive correlations (r= 0.46 to 0.76, p=0.046 to 0.001) between changes over time in depression symptoms and changes over time in both insomnia and PTSD symptoms in the CBT-D group. There were no such correlations in the control group. Participants reported higher therapeutic alliance (p=0.0001) and overall mobile application user rating (p=0.036) for the CBT-D app than for the control education app. More favorable therapeutic alliance and mobile application user ratings correlated significantly with greater improvements in depression symptoms in the CBT-D group (r=0.53 to 0.66, p=0.02 to 0.002). However, dropout was substantial (40/59 in the CBT-D group and 31/54 in the control group) and results may not be fully representative of the population at risk.

**Conclusions:** There remains a major unmet need for widely available, low-cost, effective interventions for depression symptoms in the context of mild TBI. The challenges of designing and rigorously assessing the effects such interventions remain formidable. However, the lessons learned from this study and indications of benefit in a highly relevant secondary outcome measure (insomnia) suggest that remote app-based interventions remain therapeutically viable.

**Trial Registration:** clinicaltrials.gov NCT05147506, submitted 20 July 2021, posted 7 December 2021.

## INTRODUCTION

TBI has become a frequent diagnosis among United States (US) military personnel ^1^. Since 2000, over 509,000 service members have been diagnosed with a TBI, and approximately 82% of these injuries are classified as concussion, also known as mild TBI (mTBI) (https://health.mil/Military-Health-Topics/Centers-of-Excellence/Traumatic-Brain-Injury-Center-of-Excellence/DOD-TBI-Worldwide-Numbers, accessed March 10, 2025). Depression is one of the most common neuropsychiatric sequelae of TBI ^2,3^. In military personnel, rates of depression, post-traumatic stress disorder (PTSD), and suicidal ideation increase following mTBI and almost double with multiple head injuries ^4^. In a survey of over 2,500 service members, it was found that 15% of returning Operation Enduring Freedom and Operation Iraqi Freedom service members had experienced a head trauma resulting in mTBI, and 13% of these service members reported significant symptoms of depression ^5^. Comparatively, only 6% of service members who experienced a different type of injury and 3% of service members with no injury reported similar depressive symptoms. One study found that 89% of Veterans diagnosed with mTBI also had a comorbid psychiatric diagnosis ^6^, much higher than 34% found in a civilian population with mTBI ^7^, which suggests that there are different variables affecting each population. These findings indicate that developing effective interventions for service members with depression and a history of mTBI should be considered a priority.

Additionally, the circumstances associated with the head trauma for service members may trigger moral injury, which can in turn worsen depression. Moral injury is the psychological distress, lasting emotional pain, and potentially debilitating spiritual/social consequences that can result from perpetrating, failing to prevent, bearing witness to, or learning about acts that deeply transgress one’s own deeply held moral beliefs and expectations. Moral injury can be thought of as a deep ethical wound ^8^ (https://www.ptsd.va.gov/professional/treat/cooccurring/moral_injury.asp, assessed January 27, 2026) There remains a stigma for service members when it comes to seeking mental health care and a reluctance to attribute difficulty functioning to mental health issues ^9^ ^10^. Service members and veterans often underutilize, or do not utilize, mental health care ^11^ ^12^. The stigma intertwined with mental health care in the military continues to contribute to underutilization and premature treatment dropout, ensuring this subset of warfighters does not receive sufficient mental health services ^10,13^.

In-person cognitive-behavioral therapy (CBT) is one of the most comprehensively researched forms of psychotherapy ^14^. CBT’s effectiveness in treating depressive symptoms is well documented^15,16^ ^17^. CBT is highly effective for depression versus no-treatment, wait-list, or placebo controls ^14^. Compared with supportive therapy, CBT was found to be more effective and showed greater improvements in patients’ mood and disability ^18^. Long-term, CBT had lasting effects with a 22% reduction in relapse and stable improvement at 12- and 24-months post-treatment ^19^. Cognitive therapies including CBT can be at least as effective as or more effective than antidepressant medications ^20^.

Computer-based CBT treatments for depression began in the early 1980s ^21^ ^22^. Since then, dozens of studies have found that internet and computer-based programs can reduce symptoms of depression ^23^ ^24^ ^25^ ^26^. Research has also shown that automated support in web-based interventions for depression can have similar outcomes to in-person interaction ^27^.

A systematic review of publications examining mobile applications based on CBT principles found that all eight included studies were effective to various degrees in reducing mental health symptoms, especially depressive symptoms^28^. Furthermore, using mobile applications significantly reduced depressive symptoms compared to multiple control conditions ^29^. Of note, smartphone-based interventions produced greater benefits for patients with mild to moderate depression, similar to findings with computerized CBT ^30^. Neither smartphone-based interventions nor computerized CBT were as effective for patients with severe depression. In general, CBT has also been found to be effective in the treatment of mTBI-related symptoms ^31^, and appears to have the best preliminary evidence of any psychotherapeutic approach for treating depressive symptoms following a mTBI ^32^. However, a 2018 review of smartphone applications for people with TBI found 70 applications for TBI: none of which were based on principles of CBT ^33^.

In 2019, we undertook an effort to develop a new mobile application to deliver CBT for depression designed for military service members with a history of mTBI called *Mobile <u>A</u>pplication to <u>C</u>ombat <u>D</u>epression and <u>C</u>oncussion (ACDC).* To our knowledge, ACDC is among the first CBT mobile applications developed to counteract depressive symptoms in service members and veterans with a history of mTBI. This intervention was based on the *Cognitive-Behavioral Therapy – Depression* (CBT-D) therapist manual ^34^ and *Cognitive-Behavioral Therapy – TBI* (CBT-TBI) therapist and patient manuals ^35^ ^36^. The CBT-D manual, developed by the US Department of Veterans Affairs (VA), closely follows the original CBT concepts with minor modifications to make the manual specific to military service members. The CBT-TBI manual was adapted from the structured telephone care management and CBT protocol created by the *Life Improvement Following Traumatic Brain Injury* research team ^37^. Development of the new ACDC mobile application also drew upon a previously developed scientific framework for behavioral interventions delivered via the internet. This framework was originally used to develop a web-based CBT program for insomnia, *Sleep Healthy Using the Internet (SHUTi).* SHUTi has been shown to produce similar efficacy to traditional CBT for insomnia: significant reductions in insomnia severity ^38^. SHUTi is also effective in reducing other related outcomes such as quality of life and fatigue ^39^, and psychological outcomes such as depression and anxiety ^40^. The similarities between internet-based and mobile application-based interventions allowed the development team to adapt this model for the ACDC application, ensuring the scientific integrity of the intervention, while retaining the flexibility to adapt the program to people with TBI.

Our purpose in creating the ACDC mobile application was to provide a tool that could reduce depressive symptoms in adults with TBI that is not dependent on seeing a provider. With this application, our goal was to fill in the gaps where in-person therapy is not an option, rather than reducing person-to-person interaction for people able to receive in-person treatment. This represents a major unmet medical need.

Our hypothesis was that depression symptoms in military service members and veterans with history of mTBI would improve more after completion of the ACDC mobile app than in a control group. To test this hypothesis, we designed and implemented a fully remote randomized controlled trial. We considered several options for the control group including wait-list, no intervention, and an educational mobile app. We opted for the latter, and designed a separate educational mobile application. The control application was considered “neutral” because it contains educational lessons about concussions and depression, but does not include the therapeutic components of CBT that are in ACDC. The control application was developed concurrently with the ACDC application, and used the same interface and designs to maintain consistency between groups. We also hypothesized that key secondary outcome measures assessing post-traumatic stress symptoms, insomnia, and TBI-related quality of life would similarly improve more after completion of the ACDC mobile app than in the control group.

## METHODS

### Intervention Development

The development of the ACDC intervention began in 2018 and was completed in 2021. With input from veterans and subject matter experts in CBT for patients with TBI, the development team designed a CBT mobile application that combined the most important aspects from the CBT-D therapist manual ^41^ and CBT-TBI manuals ^35,36^ to develop a novel intervention built specifically for service members and veterans with depressive symptoms and a history of mTBI. In order to properly serve this population, the development team followed the 2016 Americans with Disabilities Act compliance guidelines for websites and mobile applications (e.g., enhanced multimedia features such as audio descriptions for images as well as text captions and transcripts of sessions). To assist users who are unfamiliar with using mobile application interventions, the team filmed an introductory “How to Use this App” video that users could watch before engaging with the application.

The ACDC intervention consists of 12 weekly lessons that followed the course of CBT-TBI laid out by the *Life Improvement Following Traumatic Brain Injury* team. In-person CBT-TBI sessions typically last 30 to 60 minutes ^35^. The lessons in our mobile intervention were shorter, lasting 15 to 20 minutes for two main reasons: (1) there is no therapist, therefore the user engages at their own rate, and (2) to minimize the amount of new information the user gets each week. Shorter lessons over a longer period of time were intended to accommodate several post-concussion symptoms including impaired attention and concentration, as well as impaired learning and recall. Key features and goals of the CBT lessons included stigma minimization, personal engagement, and 1-2 hours per week of personal challenge homework assignments. The specific lessons included the following:

1. Understanding Depression, including mood tracking
2. Getting Started, including personal experiments
3. Personal Experiments, including pleasant events scheduling
4. Continuing Your Personal Experiments, including internal barriers to personal experiments
5. Adding It Up & Keeping It Going, including an activity calendar
6. Stay Active – Inner Barriers including how to get over internal roadblocks
7. Standing at a Distance, including learning to watch yourself think
8. Negative Focus - Ignoring the Positivity. Including common signs of negative thinking
9. Challenging Negative Thoughts including thought balancing
10. Practicing Thought Balancing, including core belief modification
11. Self-Care, including how to create a self-care plan
12. The Long Run, including self-care plan, thought journal, and pleasant event scheduling

The complete outline of the CBT lessons is detailed in the Supplementary Information.

The ACDC app was designed to facilitate development of therapeutic alliance using human avatars described as “coaches,” to attempt to optimize efficacy ^42^. The ACDC app included audiovisual representations using actors playing the roles of people intended to be individuals with whom the patients can identify. The use of the coaches targeted three domains: First, to emotionally engage the patient on a reciprocal basis. “Reciprocal” connotes interaction between the patient and the app to bring about changes in the attitudes and behaviors of the patient. Changes in attitudes and behaviors are, in part based on the patient’s self-identification with the coach. To the degree that the patient identifies with the coach and his or her experience, changes in attitudes and behaviors are more likely. Second, to present information for the patient to consider with which the patient can personally identify, constrained within the CBT-D curriculum. Third, to encourage the patient to undertake tasks requiring changes in behavior, attitude and self-perception in order to overcome the resistance to change that is normally encountered.

The ACDC coaches were written as individuals with separate personalities and military/life experiences to try to relate to as many users as possible. These characters were modeled on amalgams of real service members and veterans who have lived through traumatic experiences and depression but were able to improve their mood and other symptoms using CBT. None of the information provided referred specifically to real people, and there were no direct or indirect identifiers. See **Suppl. Table 1, and Suppl. Fig. 1** for additional background on this approach, script examples, and brief descriptions of each coach. To make the coaches as realistic as possible, the development team consulted with and received feedback from veterans and included veteran scriptwriters.

The ACDC app and the control app were produced by Blue Whale Apps (Fairfax, VA) with versions for both iOS and Android operating systems. The apps were produced for a one-time payment to the developer, with no subsequent charges for use of the apps after development. The framework of the app is provided as **Suppl. Fig. 2**. Access to the apps was provided via links provided by the study team. Production and troubleshooting required 15 months. The ACDC app is currently available upon request from the authors at no cost.

### Randomized Controlled Trial

We designed and executed a remote, blinded, parallel group, randomized controlled trial of the ACDC app vs the education control app. Initially the sample size was intended to be 200, with 1:1 randomization. The trial was designed to have 90% power to detect a Cohen’s d effect size of >0.6 with a p value <0.05 using a 2-sided Student t-test with 100 randomized participants per group and 60 completers per group (anticipated dropout rate of 40%). An effect size of 0.6 translates to a clinically meaningful 3-point difference between groups in reduction in Patient Health Questionnaire-9 score, the primary outcome measure, assuming a standard deviation of five points.

The initial inclusion criteria were 1) age 18-65 years, which was later relaxed to age 18-70 years, 2) able to provide informed consent; no surrogate consent was allowed, 3) current or former member of the US military; this criterion was later relaxed to allow civilian participation, 4) have a history of one or more mTBIs from any cause and in any context ≥ six months prior to enrollment, with no upper limit in number of mTBIs or time from injury to participation. The definition of mTBI was concordant with the DoD/VA guidance – TBI diagnoses were based on self-report, review of medical records and imaging were not required. 5) Baseline PHQ-9 score of ≥ 5 to 27, indicative of mild or greater depressive symptoms, 6) ownership of or reliable access to a smartphone with a data plan or internet connecting capabilities for the anticipated 16-week study duration. 7) No substantive changes in pharmacological or non-pharmacological treatment for depression within the three months prior to trial enrollment, 8) No active psychotic or bipolar symptoms, 9) No other considerations that, in the opinion of the investigators, may adversely affect patient safety, participation, or scientific validity of the data being collected (e.g., active plan or intent for suicide, terminal illness, unreliable communication). Later, the criteria were expanded to include the requirement for physical location in the United States or US-held territories after one participant was discovered to have been participating in Canada, where US Institutional Review Board (IRB) jurisdiction does not apply. No alterations in participants concomitant care were mandated during the trial period.

The study was approved by the USUHS IRB initially on 20 July 2022. The study was considered to be greater than minimal risk. The ACDC app was considered to be a nonsignificant risk device and met the abbreviated Investigational Device Exemption requirements. The study was registered on clinicaltrials.gov as NCT05147506, first submitted 20 July 2021 and first posted 7 December 2021. There were five internal monitoring visits, conducted by the clinical trials unit of the Center for Neuroscience and Regenerative Medicine/Military Traumatic Brain Injury Initiative. The name of the organization changed in 2023 but the monitoring protocols did not change. The study adhered to CONSORT Guidelines (for CONSORT Checklist, see **Supplemental Information**)

Participant recruitment involved multiple outreach methods. These included advertisements in the Washington DC public transit metro, contacting individuals registered in the TBI Research Opportunities and Outreach for Participation in Studies (TROOPS) referral program (https://mtbi2.usuhs.edu/join/study-recruitment), posting study flyers at military treatment facilities and other relevant locations, Google ads, physician referrals from TBI specialists and behavioral health providers in the military health system, advertising through Concussion Alliance (https://www.concussionalliance.org/), advertising though LoveYourBrain (https://www.loveyourbrain.com/), social media posts, and word-of-mouth referrals. The TROOPS registry and the metro ads were the most effective methods (**Fig. 1**).

**Fig. 1:**
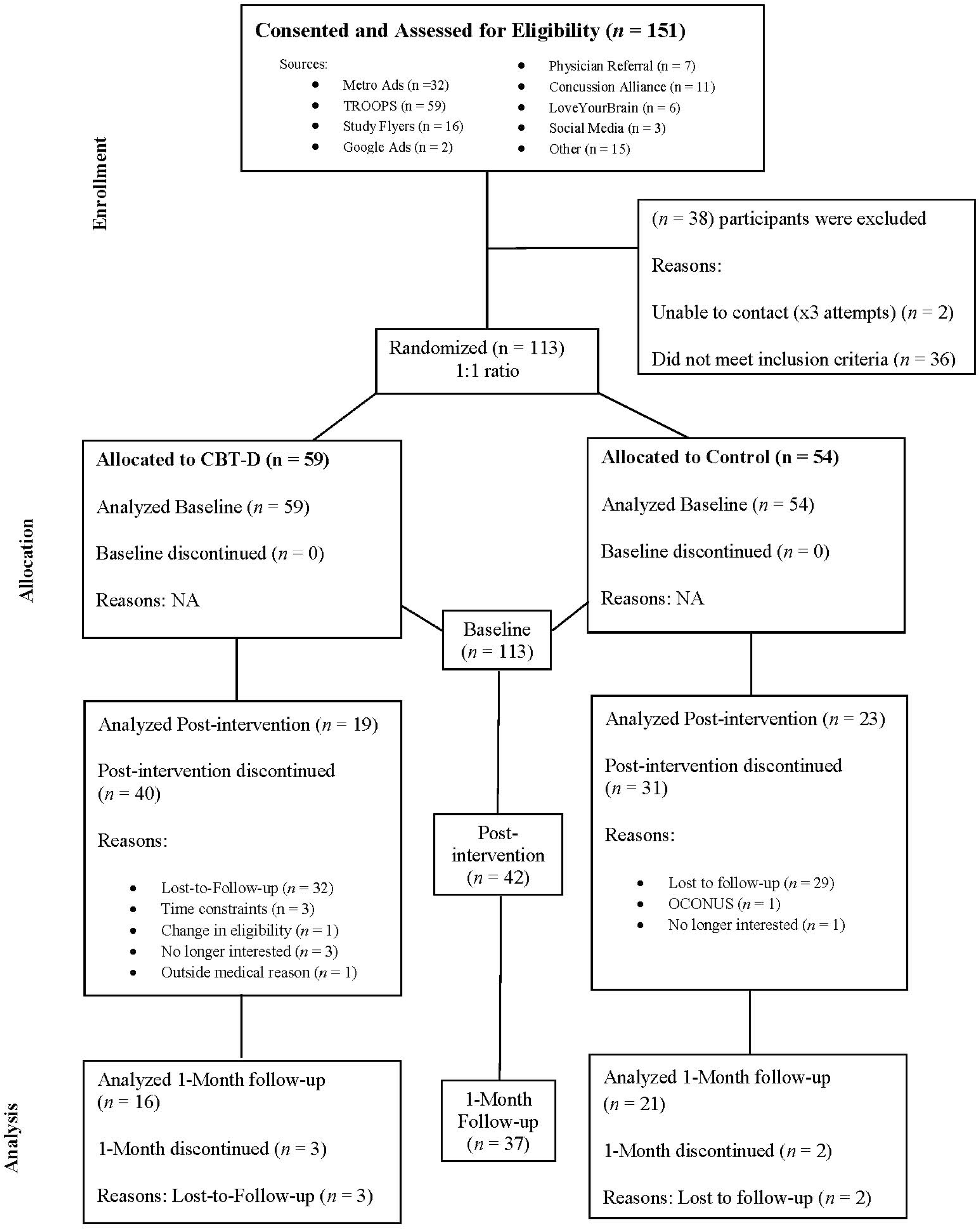
CONSORT Diagram showing flow of participants through enrollment, randomized allocation, and analysis.

The primary outcome measure was the Patient Health Questionnaire 9 for Depression (PHQ-9). The PHQ-9 is a self-report assessment for depressive symptoms ^43^ and has been validated in the military population^44^. Each item is scored on a scale of 0 to 3 providing a summary score that ranges from 0 to 27, with higher scores representing more severe depression. On the PHQ-9 a total score <5 is considered minimal depression, and scores of 5 to 9, 10 to 14, 15 to 19, and 20 to 27 are considered mild, moderate, moderately severe, and severe depression, respectively. The PHQ-9 has been shown to have satisfactory sensitivity and specificity ^43,45^ to MDD. The PHQ-9 is also a gauge of minimal clinically significant changes for depressive symptoms ^46^. A five-point reduction in PHQ-9 score is usually considered the standard definition of a clinically significant improvement for the PHQ-9 ^43^. Of note, this trial was designed to be powered to assess a three-point reduction, a smaller difference. The PHQ-9 takes approximately three to five minutes to complete. For trial eligibility the PHQ-9 was administered via telephone by trained study personnel^47^. For the baseline, 12-week post-intervention assessment, and 16-week follow-up assessment, the PHQ-9 was administered electronically ^48^. The prespecified primary outcome measure was the change in PHQ-9 from baseline to the 12-week post intervention assessment.

There were three prespecified clinical secondary outcome measures, administered electronically at baseline, 12-week post-intervention assessment, and 16-week follow-up assessment:

- Traumatic Brain Injury Quality of Life Scale (TBI-QOL): The TBI-QOL is a self-report questionnaire composed of 22 item banks ^49^. It was developed to measure multiple domains of life for patients living with TBI, including physical, mental, cognitive, and social. The TBI-QOL takes approximately 30 minutes to complete.
- PTSD Checklist for DSM-5 (PCL 5): The PCL 5 is a self-report questionnaire designed to assess symptoms of PTSD ^50^. The PCL-5 takes approximately 5-10 minutes to complete. The PCL-5 has been administered electronically ^51^.
- Insomnia Severity Index (ISI): The ISI is a self-report questionnaire designed to assess the presence and severity of primary insomnia sleep disorder and is empirically-validated in general and military populations ^52^ ^53^ ^54^. An ISI score of approximately 11 or greater has been demonstrated to be an appropriate cutoff for confirming the presence of mild clinical insomnia ^53^ ^55^. The ISI takes approximately three to five minutes to complete and has been validated for use over the Internet.

There were four additional self-report measures collected electronically, which were designed to assess correlates of response to the CBT-D intervention:

- HEXACO Personality Inventory – Revised (HEXACO-PI-R): The HEXACO-PI-R is a 100-item personality inventory that assesses six domains of personality: Honesty-Humility, Emotionality, eXtraversion, Agreeableness, Conscientiousness, and Openness to experience ^56^. It takes approximately 20 to 30 minutes to complete. HEXACO was collected at baseline.
- Credibility and Expectancy Questionnaire (CEQ): The CEQ is a six-item assessment of the patient’s beliefs and expectancies regarding the intervention ^57^. The questionnaire can be adapted for any program.
- User Version of the Mobile Application Rating Scale (uMARS): Adapted from the Mobile Application Rating Scale (MARS), the uMARS is a 26-item self-report questionnaire designed to assess the quality, functionality, and aesthetics of a mobile application and its information^58^. There is an additional open-text question for further comments on the users’ opinions of the application. The uMARS was assessed after completion of the intervention.
- Mobile Agnew Relationship Measure (mARM): The mARM is an adaptation of a valid and reliable assessment of therapeutic alliance in face-to-face therapy ^59^. The 25-item questionnaire has been adjusted for use with digital health interventions, specifically for mental health. It takes approximately 10 minutes to complete. The mARM was assessed after completion of the intervention.
- Blinding Questionnaire: At the end of the intervention period (Week 12) participants were asked which group they thought they were assigned to in order to measure the efficacy of the blinding.

Compensation was offered for successfully completing study measures and questionnaires (up to $50 total for assessments at Week 12 and Week 16).

### Statistical Analyses

All data were analyzed after completion of the study; there were no interim analyses. The primary outcome measure was the difference in the mean changes in PHQ-9 total score between baseline and 12 weeks in the active group relative to the control group. Analyses were based on intention-to-treat as it pertains to participants’ original group assignment. The distributions of the groups were examined using plots for skewedness, and Shapiro-Wilk tests of the normality assumption were conducted. These did not reveal deviations from normal distributions. The mean change in PHQ-9 over time was analyzed using a two-sided t-test in the two groups with α-level of 0.05. A separate analysis evaluated the difference in mean change in PHQ-9 total score between baseline and 16 weeks-i.e. one-month follow-up after treatment-in the active group relative to the control group. Similar between group analyses were performed for the secondary outcome measures. Linear mixed-effects models were used to assess changes in outcomes measures over time, differences between groups, and group x time interactions. A random effect intercept for each participant was included to account for repeated measures over time in individuals. Statistical results have been presented without correction for multiple comparisons. For the three secondary analyses assessed at two time points, conservative Bonferroni correction for multiple comparisons would require p<0.05/6= 0.0083 for statistical significance. Missing data was not imputed. Correlations were assessed using Pearson product moment correlations or Spearman Rank correlations depending on the normal distribution of the residuals. Correlation analyses were considered exploratory and were not corrected for multiple comparisons.

### Data Sharing

The MTBI2 Informatics Core will transfer the identifier-free research records to be stored in the MTBI2 and Federal Interagency Traumatic Brain Injury Research (FITBIR) data repositories. All elements of protected health information and personal identifying information will be removed prior to the sharing of data with the Data Repository and the FITBIR database. Data in these repositories will be open for access to qualified researchers who have requested access to the data. The ACDC app is available by request to the authors.

## RESULTS

Enrollment and follow-up completion were modest. The study team screened 151 potential participants from August 10, 2022 through June 3, 2024. Of these, 113 were randomized to CBT-D vs. control, and 38 were excluded (**Fig. 1**). There was substantial loss-to-follow-up after randomization, with 19 of 59 participants in the CBT-D group and 23 of 54 participants in the control group completing the 12-week post-intervention primary outcome assessment. An additional three participants in the CBT-D group and two participants in the control group were lost to follow-up before the 16-week (one-month post-intervention) assessment. The demographics of those randomized to the two arms were similar (**Table 1**), including mean ages of late 40’s, approximately half women, with a predominance of self-reported “White” race and “Non-Hispanic or Latino” ethnicity. Approximately half had a reported education level of “graduate degree.” Current Active Duty Service Members, Veterans, and civilians were enrolled. The demographics of those who completed the primary outcome assessment were similar to those who were randomized but did not complete the primary outcome assessment (**Suppl. Tables 2-3**), with two notable exceptions: First, self-reported “Black or African American” participants were less likely to complete the study than self-reported “White” participants (1/8 vs. 39/52, chi-squared 12.2, p=0.0005). Second, self-reported “Hispanic or Latino” participants were less likely to complete the study than self-reported “Non-Hispanic or Latino” participants (1/14 vs. 42/54, chi-squared 23.8, p=0.0001). One protocol deviation required amendment and re-approval; one participant was discovered to have been participating in Canada, where US IRB jurisdiction does not apply, and the study inclusion criteria were modified to add the requirement for physical presence in the United States. Partway through the study, the inclusion criteria were modified to allow civilian participation and participants up to age 70 because of slow enrollment. The study was stopped by the sponsor before the prespecified enrollment numbers were achieved because the period of performance came to an end and funding was no longer available.

**Table 1:** Demographics for participants who completed the primary outcome measure.

|  | Allocated to CBT-D (n = 19) | Allocated to Control (n = 23) |
| --- | --- | --- |
| Self-reported Age, years, mean (SD) | 48.1 (12.9) | 45.7 (11.1) |
| Self-reported Gender, n (%) |  |  |
| Female | 9 (47.4%) | 12 (52.2%) |
| Other | 0 (0.0%) | 1 ( 4.3%) |
| Self-reported Race, n (%) |  |  |
| White | 17 (89.5%) | 21 (91.3%) |
| Black or African American | 0 ( 0%) | 1 ( 4.3%) |
| Asian | 0 ( 0%) | 1 ( 4.3%) |
| American Indian or Alaskan Native | 1 (5.2%) | 0 ( 0%) |
| Other (includes multiple races) | 1 (5.2%) | 0 ( 0%) |
| No Response | 0 ( 0%) | 0 ( 0%) |
| Self-reported Ethnicity, n (%) |  |  |
| Hispanic or Latino | 0 (0.0%) | 1 ( 4.3%) |
| Non-Hispanic or Latino | 19 (100.0%) | 22 (95.7%) |
| No Response | 0 (0.0%) | 0 (0.0%) |
| Self-reported Educational level, n (%) |  |  |
| Partial degree | 0 (0.0%) | 0 (0.0%) |
| High school degree or less | 1 (5.3%) | 2 ( 8.7%) |
| Associates degree | 4 (21.1%) | 4 (17.4%) |
| Some college or college degree | 3 (15.8%) | 9 (39.1%) |
| Graduate Degree | 11 (57.9%) | 8 (34.4%) |
| Self-reported Military Status |  |  |
| Active Duty/Full-time | 2 (10.5%) | 6 (26.1%) |
| Civilian | 9 (47.4%) | 8 (34.8%) |
| Retired | 3 (15.8%) | 2 ( 8.7%) |
| Separated/Discharged | 4 (21.1%) | 3 (13.0%) |
| Profile/Limited | 0 (0.0%) | 1 ( 4.3%) |
| No Response | 1 (5.3%) | 3 (13.0%) |
| Self-reported Employment Status |  |  |
| Full-time | 6 (31.6%) | 12 (52.2%) |
| Part-time | 1 (5.3%) | 3 (13.0%) |
| Self-employed | 0 (0.0%) | 1 ( 4.3%) |
| Unpaid (retired, student, disabled) | 11 (57.9%) | 4 (17.4%) |
| Unemployed | 1 (5.3%) | 2 ( 8.7%) |
| No Response | 0 (0.0%) | 1 ( 4.3%) |
| Self-reported Military Branch |  |  |
| Air Force | 2 (10.5%) | 1 ( 4.3%) |
| Army | 2 (10.5%) | 7 (30.4%) |
| Marine Corps | 0 (0.0%) | 2 ( 8.7%) |
| Navy | 4 (21.1%) | 2 ( 8.7%) |
| More than 1 | 1 (5.3%) | 2 ( 8.7%) |
| No Response | 10 (52.6%) | 9 (39.1%) |
| Self-reported Rank |  |  |
| Enlisted | 6 (31.6%) | 6 (26.1%) |
| Officer | 3 (15.8%) | 8 (34.8%) |
| No Response | 10 (52.6%) | 9 (39.1%) |
| Self-reported Military Occupation |  |  |
| Combat | 4 (21.1%) | 13 (56.5%) |
| Non-Combat | 8 (42.1%) | 3 (13.0%) |
| No Response | 7 (36.8%) | 7 (30.4%) |

There was no difference between the groups randomized to CBT-D vs. control in the prespecified primary outcome measure, which was between groups differences in change from baseline to post-intervention in self-reported depression symptoms, as assessed via the PHQ-9 (**Fig. 2A**). The changes in PHQ-9 scores from baseline to post-intervention were 2.0 in the CBT-I group and 3.1 in the control group. Similarly, there was no difference in PHQ-9 between groups at the 16-week follow-up assessment. Both groups improved over time and were generally in the ‘moderate’ symptom severity range. In a linear model, there was a decrease of 2.1 points from baseline to the post-intervention follow-up (p=0.005) and a decrease of 3.1 points at the 16-week follow-up assessment (p=0.0001). The differences between groups in the linear model were non-significant, as were the interactions between group and time. Likewise, there were no statistically significant differences between groups for responses to any of the individual PHQ-9 questions in post-hoc exploratory analyses (**Suppl Fig. 3**). The sample sizes (n=19 in the CBT-D group, and n=23 in the control group) were not large enough to allow meaningful statistical subgroup analyses.

**Fig. 2:**
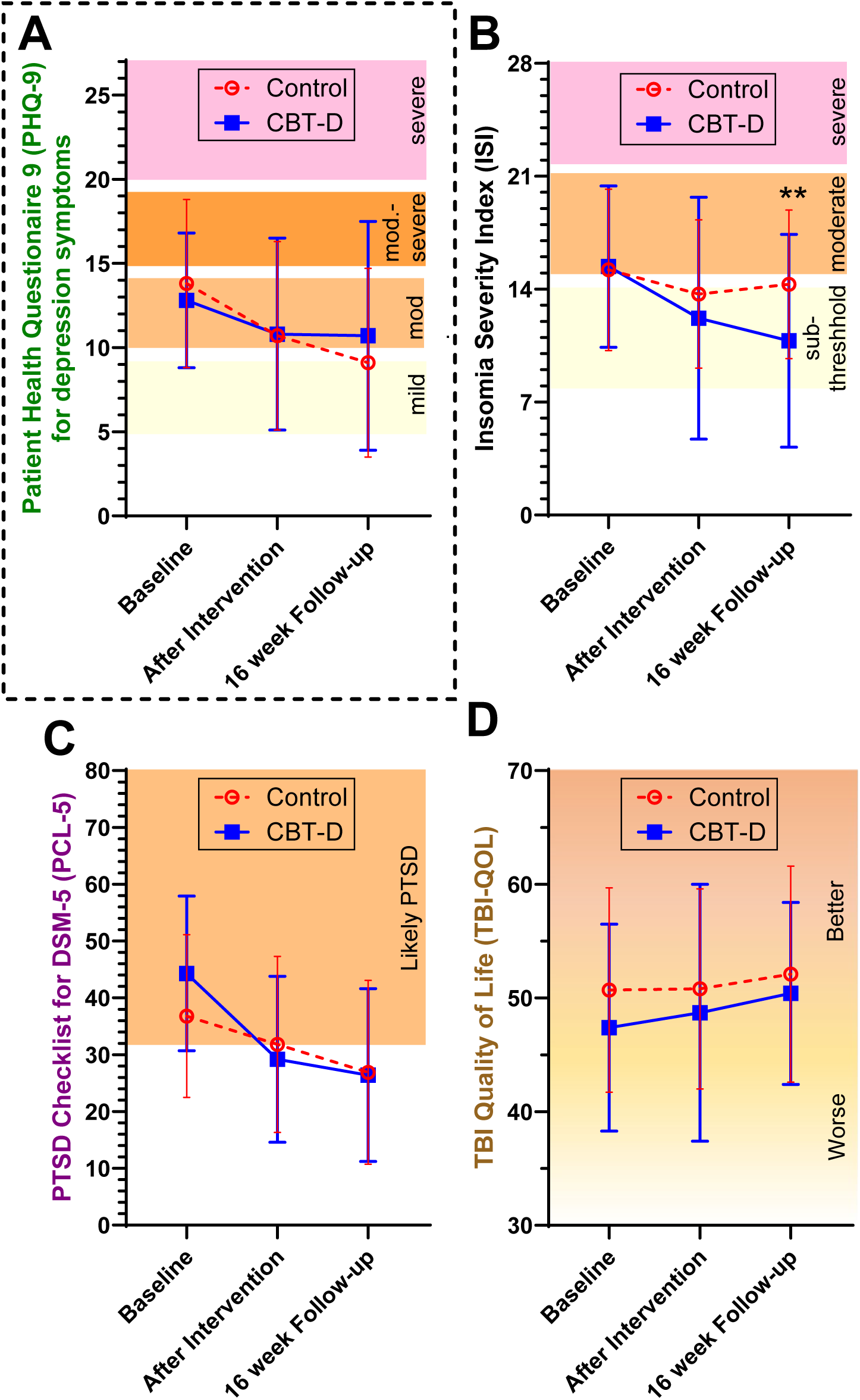
Changes over time in the primary outcome measure and key secondary outcome measures. **A.** Change over time in self-reported depression symptoms, assessed via the PHQ-9. PHQ-9 was the prespecified primary outcome measure. PHQ-9 scores range from 0 to 27, with higher scores indicating more severe depression symptoms. **B.** Change over time in self-reported insomnia, assessed via the ISI. ISI scores range from 0 to 28, with higher scores indicating more severe self-reported insomnia symptoms, ** indicates p=0.0064. **C.** Change over time in self-reported PTSD symptoms, assessed via the PCL-5. PCL-5 scores range from 0 to 80, with higher scores indicating more severe self-reported PTSD symptoms. **D.** Change over time in TBI-related quality of life, assessed via the TBI-QOL. TBI-QOL scores have a mean of 50 and a standard deviation of 10, with higher scores indicating more favorable quality of life. Error bars indicate standard errors.

There were no unexpected or serious safety concerns. There were no unexpected adverse events or serious adverse events. According to our IRB approved protocol, we tracked all reports of suicidal ideation or severe depression during the study regardless of whether they were part of the natural history of the disease, related to the intervention, or represented a change from baseline. There were 24 reports of suicidal ideation (PHQ-9 question 9 score >0) in eight individuals in the control group; 11 reports were from one individual and six reports were from another individual. There were 20 reports of suicidal ideation in 10 individuals in the treatment group; seven reports were from one individual and four reports were from another individual. The differences between groups were not significant. There were eight reports of severe depression, prespecified as PHQ-9 total score >19, with four in each group. After each report, the team followed-up directly with the participants by phone or email to ensure that an acceptable clinical safety plan was in place. All participants reported that their suicidal ideation had resolved, they were in treatment with a behavioral health provider, they were working to find a provider, or more than one of the above. The majority (30/44) of the follow-ups were completed within one day of the reports (range 0-23 days) with delays due to slow responses from participants. There were no deaths from any cause, no suicide attempts, no psychiatric hospitalizations, and no other adverse events in any of the participants of which we are aware. There were no protocol violations.

There were statistically significant benefits of the CBT-D intervention in some prespecified secondary outcome measures. Most notably, there was a statistically significant improvement in self-reported insomnia severity index (ISI) at the 16-week follow-up assessment (**Fig. 2B**). Both groups started in the sub-threshold to moderate insomnia categories; there was negligible change in the control group, but the CBT-D group moved into the subthreshold range by 16-weeks. In a linear model, the factor representing interaction between group and change over time from baseline to 16-weeks was statistically significant (Cohen’s D=0.45, 95% confidence interval 0.12-0.96, p=0.0064). This would be considered significant even after conservative correction for multiple comparisons (three prespecified secondary outcome measures x two time points). In addition, there was a more rapid improvement in self-reported PTSD symptoms (PCL-5) in the CBT-D group than in the control group (**Fig. 2C**). In a linear model, the factors representing interaction between group and changes over time from baseline to post-intervention and to 16-weeks were both statistically significant (p=0.0023 and p=0.024). However, the CBT-D group started out with higher scores at baseline and the two groups were not different in absolute scores at either of the follow-up time points. There were no statistically significant differences between groups or interactions between group and time for self-reported TBI quality of life (**Fig. 2D**).

Because of the nature of the study, it was not possible to fully blind the participants. Nonetheless, when asked whether they thought they were receiving the CBT-D intervention or the education control intervention, 15/23 guessed correctly that they were in the control group, and 6/19 guessed correctly that they were in the CBT-D group. The difference was not statistically significant (Chi square 4.96, p=0.08).

There were statistically significant correlations between changes in depression symptoms and changes in some secondary outcomes in the CBT-D group. Specifically, there was a positive correlation between change in PHQ-9 and change in PTSD after intervention (r=0.46, p=0.045, **Fig. 3A**) and at 16-week follow-up (r=0.53, p=0.046, **Fig. 3B**). These correlations were not present in the control group (**Fig. 3C-D**). Likewise, there was a positive correlation between change in PHQ-9 and change in PCL-5 after intervention (r=0.61, p=0.007, **Fig. 4A**) and at 16-week follow-up (r=0.76, p=0.001, **Fig. 4B**). These correlations were again not present in the control group (**Fig. 4C-D**). There were no correlations between changes in PHQ-9 and changes in self-reported TBI-related quality of life (TBI-QOL) in either group at either time point (**Fig. 5**). The scatter plots of these correlations indicated that there were not apparent clusters of ‘responders’ vs. ‘non-responders’ in any of the outcome measures. HEXACO and CEQ were collected but data was not included in these analyses due to lack of statistical power.

**Fig. 3:**
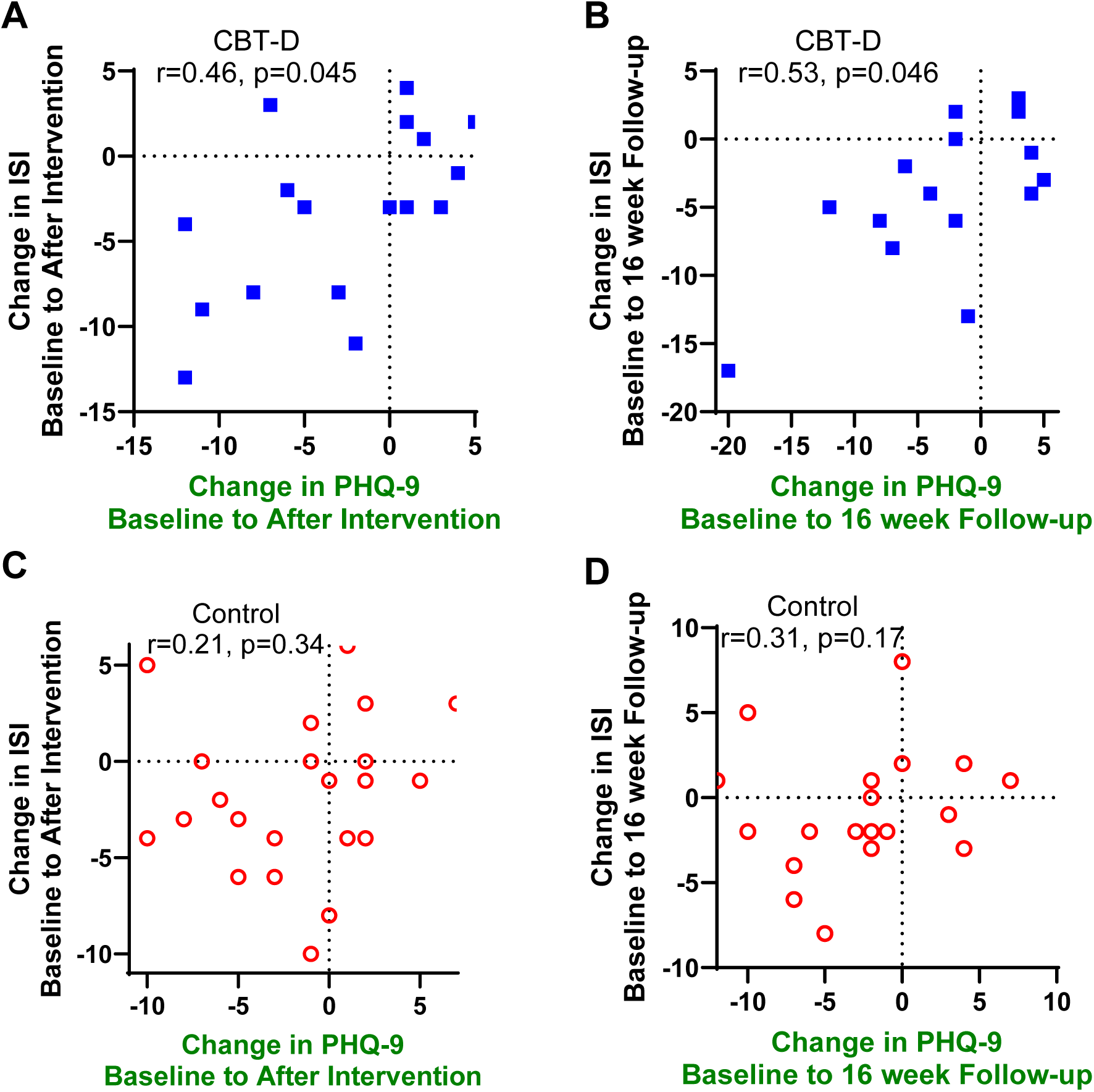
Correlations between changes in self-reported depression symptoms (PHQ-9) and changes in self-reported insomnia (ISI). **A.** Correlation between changes from baseline to after intervention in the CBT-D group. **B.** Correlation between changes from baseline to 16-week follow-up in the CBT-D group. **C.** Correlation between changes from baseline to after intervention in the control group. **D.** Correlation between changes from baseline to 16-week follow-up in the control group.

**Fig. 4:**
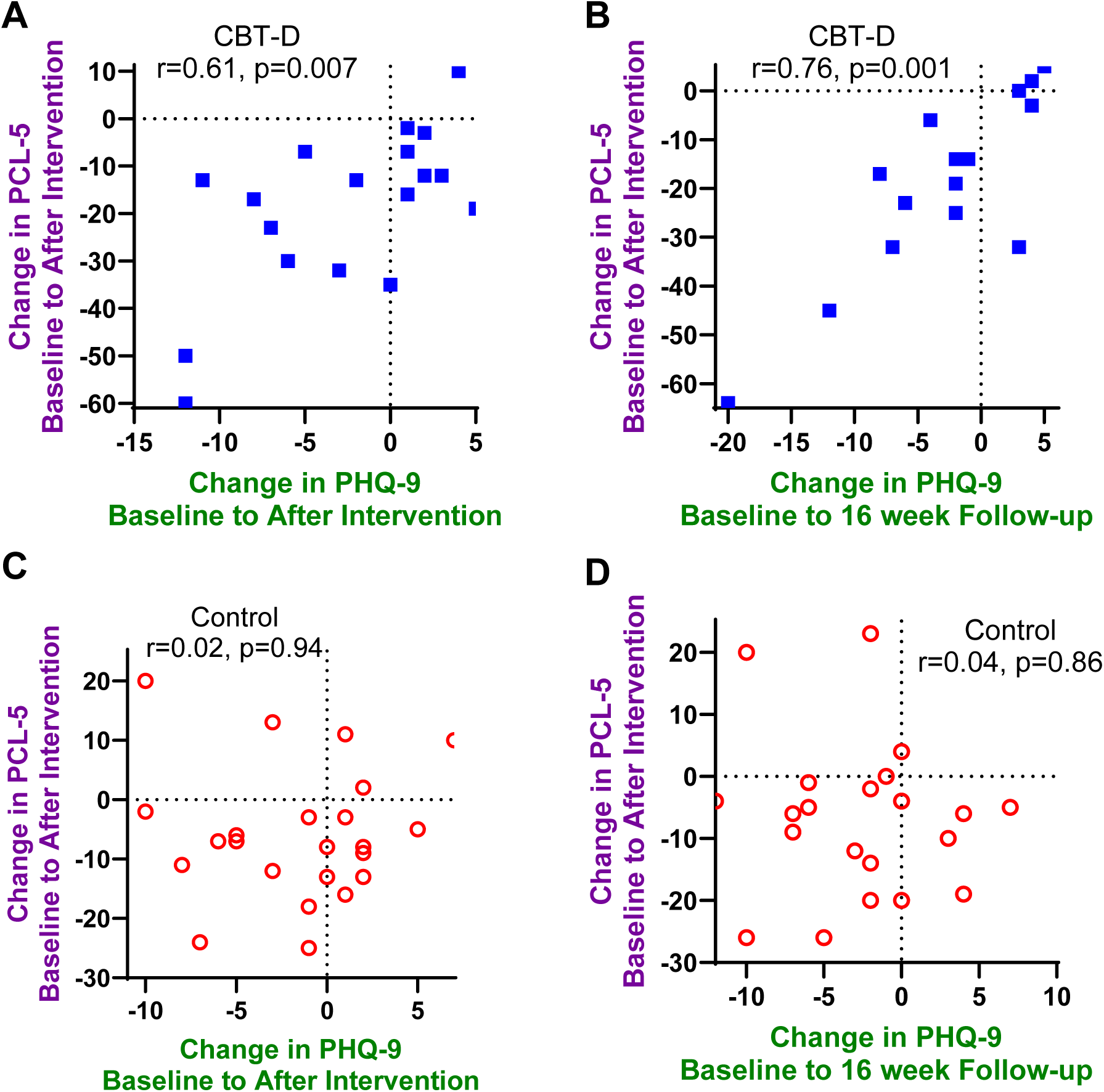
Correlations between changes in self-reported depression symptoms (PHQ-9) and changes in self-reported PTSD symptoms (PCL-5). **A.** Correlation between changes from baseline to after intervention in the CBT-D group. **B.** Correlation between changes from baseline to 16-week follow-up in the CBT-D group. **C.** Correlation between changes from baseline to after intervention in the control group. **D.** Correlation between changes from baseline to 16-week follow-up in the control group.

**Fig. 5:**
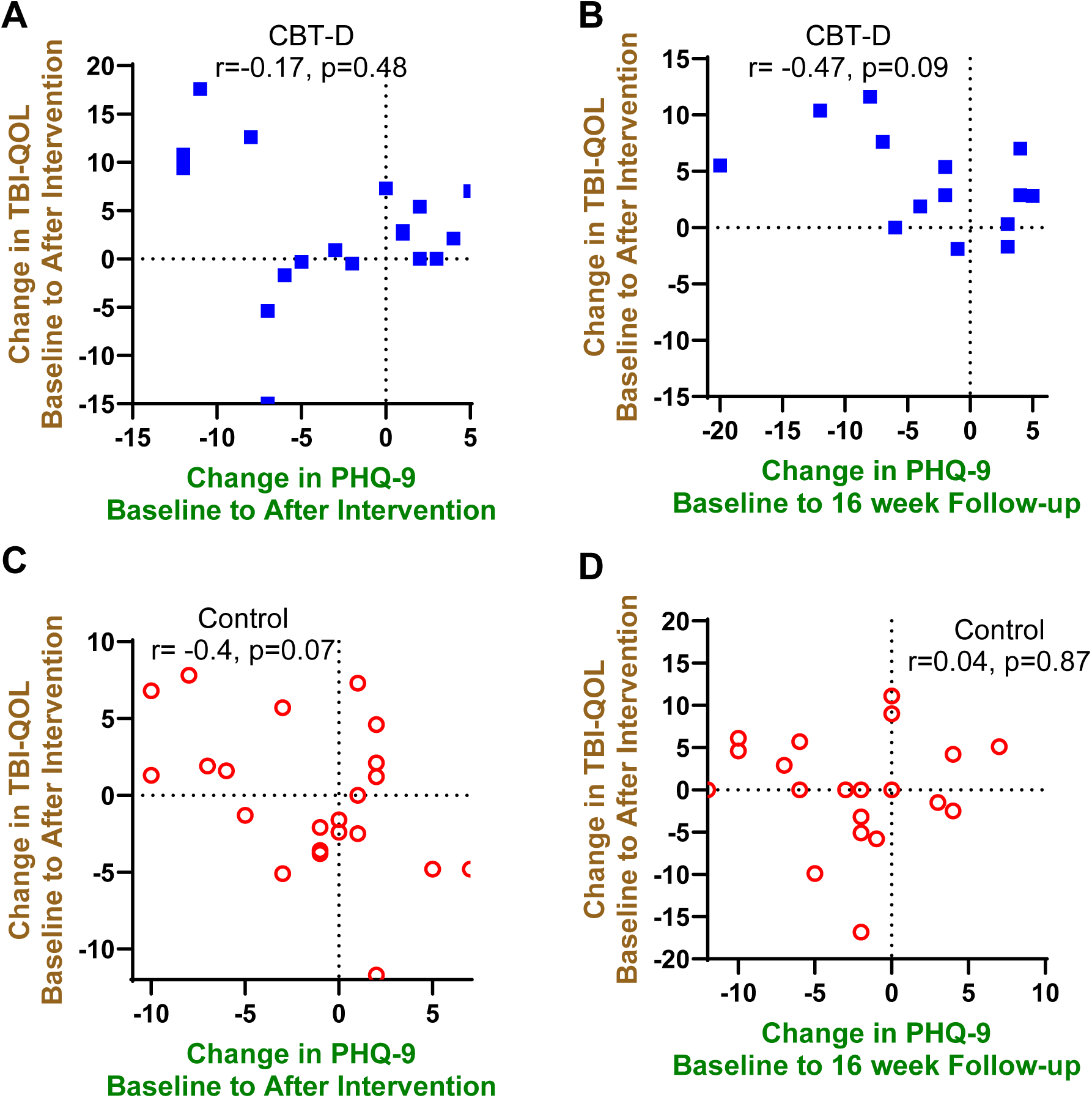
Correlations between changes in self-reported depression symptoms (PHQ-9) and changes in TBI-related quality of life (TBI-QOL). **A.** Correlation between changes from baseline to after intervention in the CBT-D group. **B.** Correlation between changes from baseline to 16-week follow-up in the CBT-D group. **C.** Correlation between changes from baseline to after intervention in the control group. **D.** Correlation between changes from baseline to 16-week follow-up in the control group.

After completing the interventions, participants generally rated the CBT-D intervention more favorably than the control educational intervention. Specifically, the CBT-D intervention was rated statistically significantly more favorably than the control intervention in terms of therapeutic alliance (p=0.0001, **Fig. 6A**) and overall mobile application rating (p=0.036, **Fig. 6B**). The CBT-D intervention was rated higher in engagement (p=0.002, **Fig. 6C**) but not in functionality (**Fig. 6D**), aesthetics (**Fig. 6E**), or information quality (**Fig. 6F**). Most ratings were in the moderately favorable range (4 out of 5), but there were a few outliers in both groups with low ratings. Of note, engagement ratings were somewhat lower (typically in the 3 out of 5 range) than functionality, aesthetics, and information quality ratings. Anecdotally, some users reported expecting more interactive apps, (e.g. artificial intelligence-based chat agents) rather than relatively static content.

**Fig. 6:**
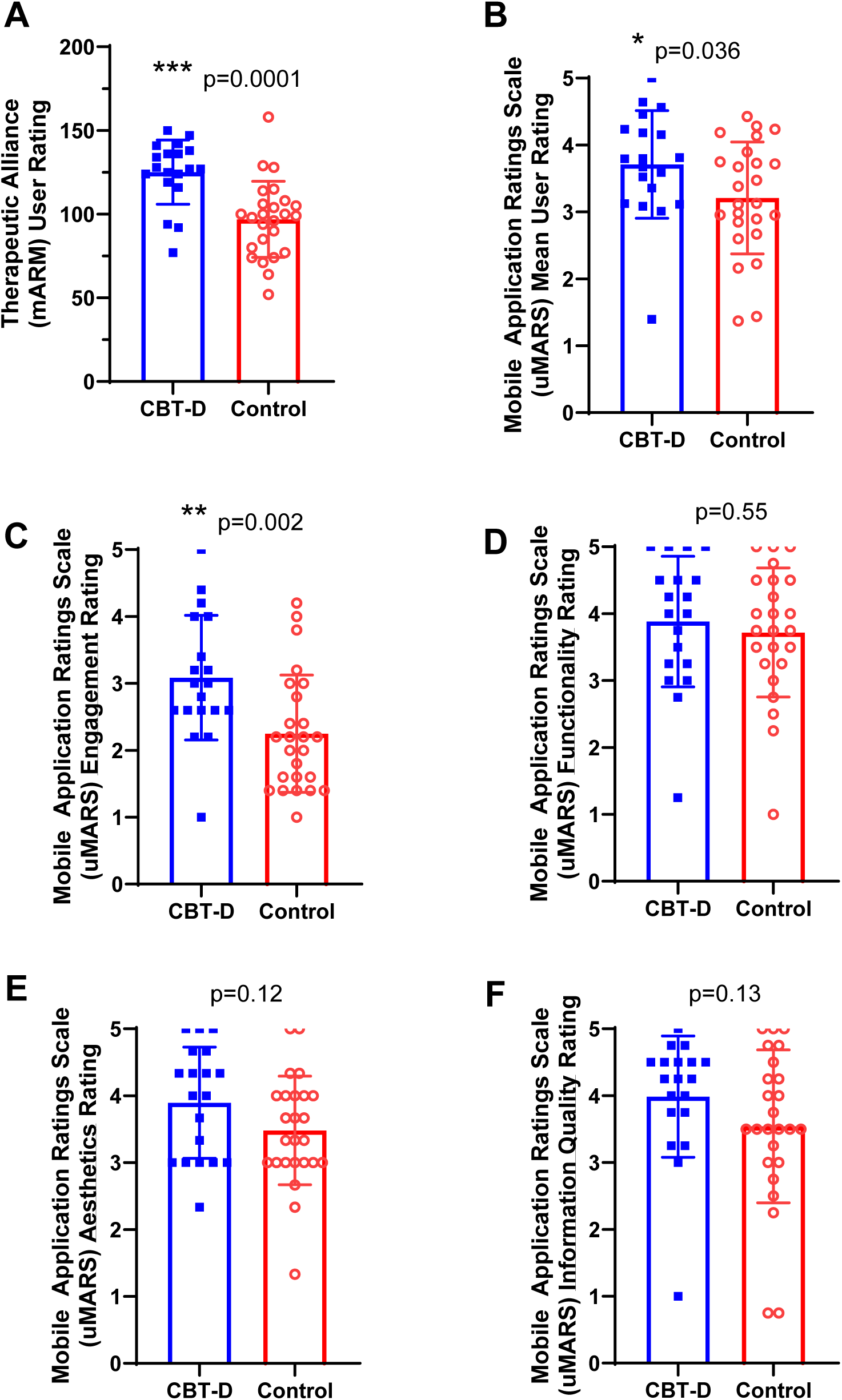
User ratings of the interventions after completing interventions. **A.** Therapeutic alliance rating using the mobile Agnew Relationship Measure (mARM). Higher mARM ratings indicate more favorable therapeutic alliance. **B.** User version of the Mobile Application Rating Scale (uMARS) mean ratings across all domains. Higher uMARS ratings indicate more favorable views. **C.** uMARS engagement ratings. **D.** uMARS functionality ratings. **E.** uMARS aesthetics rating. **F.** uMARS information quality ratings.

There were significant correlations between most of the participants’ ratings of the CBT-D intervention and its effects on depression symptoms. The therapeutic alliance ratings were higher in participants for whom depression symptoms improved most from baseline to after intervention (r= -0.63, p=0.004, **Fig. 7A**). Likewise, overall mobile application user ratings were higher in participants for whom depression symptoms improved most (r= -0.59, p=0.008, **Fig. 7B**). These correlations held for engagement ratings (r= -0.57, p=0.01, **Fig. 7C**), aesthetics ratings (r= -0.66, p=0.002, **Fig. 7E**), and information quality ratings (r= -0.53, p=0.02, **Fig. 7F**), but not for functionality ratings **(Fig. 7D**).

**Fig. 7:**
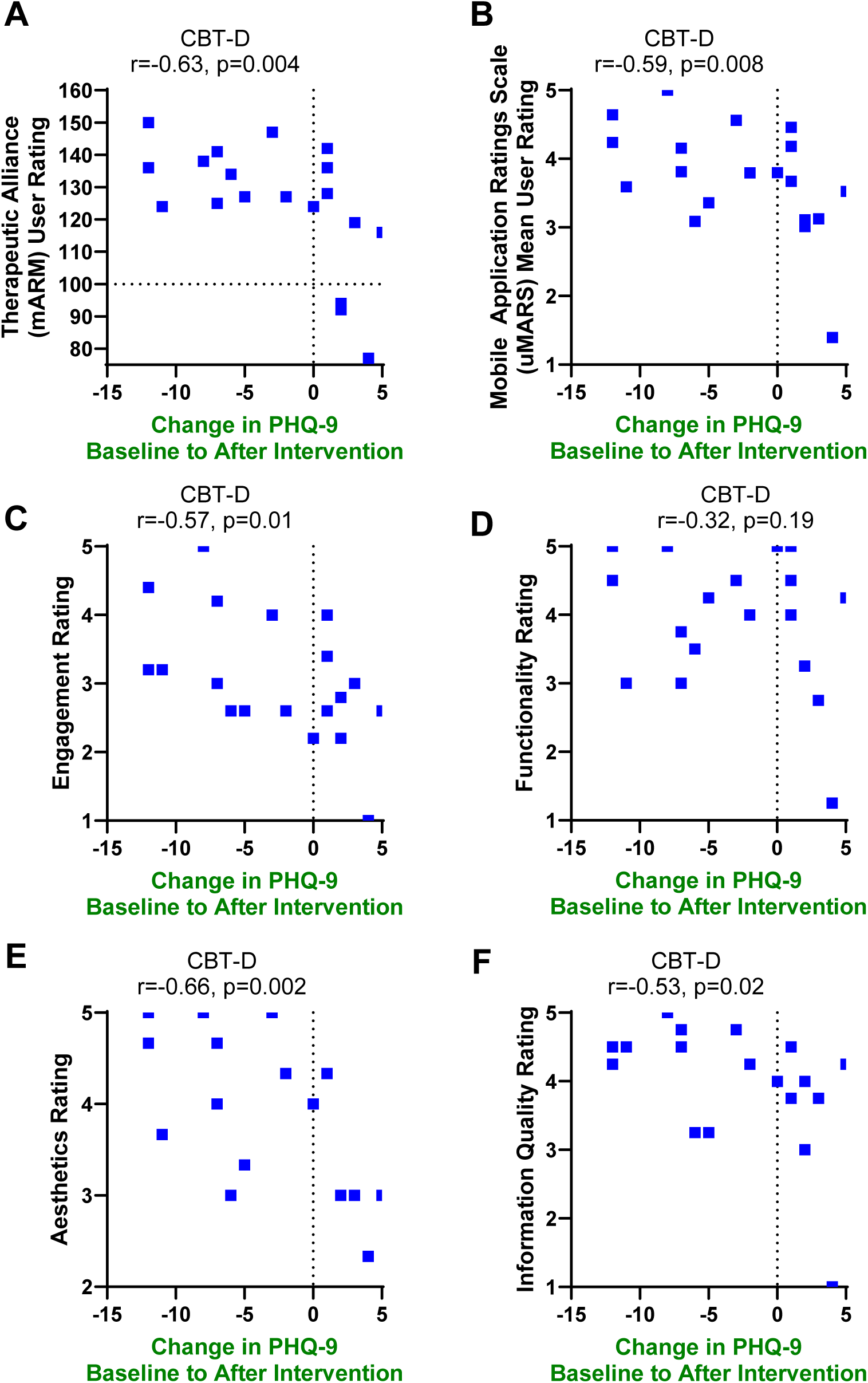
Correlations between changes in self-reported depression symptoms (PHQ-9) from baseline to after intervention vs. user ratings in the CBT-D group. **A.** Correlation vs. therapeutic alliance ratings. **B.** Correlation vs. overall mobile application ratings. **C.** Correlation vs. engagement ratings. **D.** Correlation vs. functionality ratings. **E.** Correlation vs. aesthetics ratings. **F.** Correlation vs. information quality ratings.

## DISCUSSION

In summary, although there was not a significant difference between groups in the prespecified primary outcome measure, there were indications that the CBT-D intervention had beneficial effects in clinically relevant secondary outcomes. Furthermore, there was a pattern of correlations in the participants randomized to CBT-D that indicated a spectrum of consistent positive effects not seen the participants randomized to the control intervention. There did not seem to be a clear clustering of ‘responders’ vs ‘non-responders’ based on scatter plots of the data. The modest sample sizes limited the relevance of subgroup analyses.

There are several possible explanations for why depression symptoms in both groups improved over time. Depression symptoms naturally wax and wane, and it is likely that participants are most predisposed to sign up for a research study involving candidate treatment for depression symptoms at times when the intensity of their symptoms is waxing. Thus, the improvements over time may represent the natural history of the condition. It is also possible that the education app designed to serve as a “neutral” control was more effective than we had anticipated, and both interventions should be considered “active” in this context. Finally, it is possible that the CBT-D app was not effective, and participation in either arm of the study served as a placebo. Additional studies involving a wait-list control group, longer follow-ups, and an even more rigorously neutral control intervention would be required to address these possibilities. Furthermore, given potential limitations in attention and memory in this population, delivering short lessons more frequently than once per week could be more effective than the weekly lessons used here.

Even though the CBT-D intervention was aimed at multiple aspects of depression symptoms, insomnia severity index was the most notable outcome measure that significantly changed more in the CBT-D group than in the control group. Insomnia may be especially malleable target for remote CBT-based interventions. The results reported here are concordant with the findings from our recently published randomized controlled trial of internet-based cognitive behavioral therapy for insomnia (CBT-I) in adults with TBI ^60^. In that study, ISI was the prespecified primary outcome measure and the effect size was 0.32 (95% confidence interval 0.04 to 0.70). For comparison, the effect side of the CBT-D app was 0.45, which is similar in magnitude. Another CBT-I app called “CBT-I Coach” with similar content is widely used and freely available through the US Department of Veterans Affairs in both iOS and Android formats ^61–63^.

This study had several limitations. First, the sample size was not as large as had been prespecified, thus the study lacked power to detect potentially more modest benefits. Specifically, post-hoc calculations indicated that statistical power was 0.24 to detect a 3-point difference in PHQ-9 between the CBT-D and control groups. Nearly half (9/19) of the participants in the intervention group were civilians, even though the intervention was designed primarily for military service members and veterans. We do not know whether the inclusion of civilians affected the results. The study was not adequately powered to assess subgroups in a meaningful fashion. Second, the drop-out rate was substantial, raising the question of generalizability, acceptability, and feasibility. It was notable that self-reported “Black or African American” and “Hispanic or Latino” participants differentially dropped out of the study. The reasons for drop out are not known, since the most common type of drop-out was loss-to-follow-up, which precluded gathering information about the reasons for dropping out. Future studies of this type should consider offering pre-study preference trials in tandem with a motivational interview screening approach to improve commitment to follow through with the study. A recent very successful digital therapeutics study involving cancer patients which reported that digital therapeutic engagement was comparable to in-person session attendance offers important lessons in this domain ^64^. As part of our retention efforts, we prioritized proactive communication with participants to ensure the collection of primary outcome measures. This included reaching out multiple times through various channels such as phone calls, emails, and text messages. Nonetheless, more aggressive reminders and increased compensation for participation may also be required to reduce drop-out. Third, the 100% virtual nature of the study drove the requirement to use self-reported outcome measures, rather than clinician ratings as are commonly used in clinical trials in the field. The study was not designed to quantitatively measure adherence. Participants could do homework assignments on their own without using the ACDC app, and therefore homework compliance could not be directly assessed. Likewise, the study was not designed to measure concurrent treatments. This could have affected dropouts and outcomes in those who completed participation. Fourth, the user ratings of the digital intervention were generally good, but there is clearly room for improvement. For example, ratings of ‘engagement’ were modest (mean of 3 on a 1 to 5 Likert scale), and therapeutic alliance was not rated highly by all participants. Improvements in the digital intervention could lead to better outcomes in future trials, as suggested by the statistically significant correlations between user ratings of the digital intervention and improvement in depression symptoms. We do not know whether the 12-week format or the 15-20-minute lesson length were optimal. In other structured internet and mobile-based CBT interventions, programs included five to six lessons that were 30 minutes ^65^ to 60 minutes ^66^ in length. These were completed over 3 or 6 weeks, respectively. Another program delivered 5 sessions lasting 20 to 40 minutes each, completed over 32 days ^67^. Furthermore, it is possible that the study burden could be reduced; the lengthier assessments including TBI-QOL and HEXACO could probably be eliminated without loss of meaningful information.

Studies published recently suggest that mood disorders in patients with TBI may be difficult to treat with digital therapeutics. A randomized controlled trial of a smartwatch app for stress management vs. usual care in military veterans with TBI and PTSD demonstrated benefit in some outcome measures, but no effects on self-reported depression, anxiety, or PTSD symptoms^68^. A pilot study of an evidence-based emotional regulation and problem-solving smartphone app vs conventional care in military service members with TBI also did not demonstrate benefits, though the sample size was small ^69^. Furthermore, a randomized controlled trial of an online self-management intervention vs. usual care in adolescents with recent concussion showed no overall benefit in quality of life or coping strategies ^70^. Finally, a recent randomized controlled trial of an evidence-based self-monitoring smartphone app vs. wait-list control in patients with TBI demonstrated no significant differences in self-reported anxiety and depression symptoms ^71^. A smartphone app-based mindfulness intervention for adolescents with acute concussion has been designed and is being tested in a randomized controlled trial ^72^ but results have not been presented to our knowledge. Our previous study involving internet-based CBT for insomnia was beneficial, but the effect size was smaller than was previously reported in other populations and there was no significant effect on depression symptoms ^60^.

In contrast, the efficacy of internet- and mobile-based interventions for anxiety in other contexts continues to be well-established, with more than 26 randomized controlled trials identified in a recent review ^73^. More recently, an app-based version of cognitive behavioral stress management specifically created for patients with cancer was recently shown to be effective in a randomized controlled trial using a stylistically similar health education app as a control. ^64^ ^74^. Another app designed specifically for adolescents with depression ^75^ has been completed, and positive results have been posted on clinicaltrials.gov but not yet published to our knowledge. A recent meta-analysis of 154 randomized controlled trials of internet-delivered CBT across a variety of contexts concluded that long-term efficacy was similar to short-term efficacy and that there were no significant differences between guided and self-guided internet-delivered CBT ^76^.

Since this study was conceived and the intervention was designed in 2019-2020, there has been a revolution in artificial intelligence capabilities; a digital therapeutic intervention designed now could potentially involve a much more dynamic, adaptive content by incorporating an artificial intelligence-based chat agent. In fact, some participants anecdotally reported expecting this sort of adaptive engagement. Notably, only 6/19 randomized to the CBT-D app guessed that they were receiving the CBT intervention rather than the education control, which may indicate that the ACDC app was not what they were expecting from an active CBT intervention. Qualitative approaches would be beneficial additions to future research in this domain.

## CONCLUSIONS

In conclusion, there remains a major unmet need for widely available, low-cost, effective interventions for depression symptoms in the context of TBI. The challenges of designing and rigorously assessing the effects such interventions remain formidable. However, the lessons learned from this study and indications of benefit in a highly relevant secondary outcome measure (insomnia) suggest that remote app-based interventions remain therapeutically viable.

## Data Availability

Data and materials are available from the corresponding author upon reasonable request.

## DECLARATIONS

### Abbreviations

ACDC: Mobile <u>A</u>pplication to <u>C</u>ombat <u>D</u>epression and <u>C</u>oncussion
CBT-D: Cognitive behavioral therapy for depression
CBT-I: cognitive behavioral therapy for insomnia
CBT-TBI: Cognitive-Behavioral Therapy – TBI
CEQ: Credibility and Expectancy Questionnaire
FIRBIR: Federal Interagency Traumatic Brain Injury Research
ISI: Insomnia Severity Index
mARM: Mobile Agnew Relationship Measure
mTBI: mild traumatic brain injury
PHQ-9: Patient Health Questionnaire 9 for Depression
PTSD: post-traumatic stress disorder
PCL 5: PTSD Checklist for DSM-5
SHUTi: Sleep Healthy Using the Internet
TBI: traumatic brain injury
TBI-QOL: Traumatic Brain Injury Quality of Life Scale
TROOPS: TBI Research Opportunities and Outreach for Participation in Studies
uMARS: User Version of the Mobile Application Rating Scale

### Ethics approval and consent to participate

Human studies ethics were assessed and approved by the USUHS Institutional Review Board. All participants provided written informed consent. The study adhered to the Declaration of Helsinki.

### Consent for publication

No individual participants are identified. All authors, the sponsor, and the Uniformed Services University provide consent to publish the manuscript.

### Availability of data and materials

Data and materials are available from the corresponding author upon reasonable request.

### Authors’ contributions

JBE and LM designed the app content, wrote the coach scripts, and supervised the app production. OMS and NM conducted the randomized controlled trial. MRC provided project management and oversight. AF, TJH, and DLB analyzed data. DLB conceived the project, obtained funding, assisted with safety monitoring, supervised the overall project, and wrote the first draft of the manuscript. All authors edited the drafts, and read and approved the final manuscript.

## Acknowledgements

We are grateful to the participants for their efforts. We would like to thank the following individuals for their contributions to this project: Leighton Chan, Giovanni Cizza, Kerry Dunbar, Sarah Fitzsimmons-Molina, Adele Fu, Nasreen Jahed, Taylor Knapp, Yvonne Maddox, Molly Malarkey, Dominic Nathan, Victoria Nguyen, Michael Roy, Patricia Spangler, Billy Terrell, Mark Whiting, Camille Young, Kirsten Youngren, and all of the clinicians who referred potential participants.

## References

1 Terrio, H. et al. Traumatic brain injury screening: preliminary findings in a US Army Brigade Combat Team. The Journal of head trauma rehabilitation 24, 14–23 (2009). 10.1097/HTR.0b013e31819581d8

2 Jorge, R. E. & Arciniegas, D. B. Neuropsychiatry of traumatic brain injury. The Psychiatric clinics of North America 37, xi–xv (2014). 10.1016/j.psc.2013.12.002

3 Stein, M. B. et al. Risk of Posttraumatic Stress Disorder and Major Depression in Civilian Patients After Mild Traumatic Brain Injury: A TRACK-TBI Study. JAMA psychiatry (2019). 10.1001/jamapsychiatry.2018.4288

4 Bryan, C. J. & Clemans, T. A. Repetitive traumatic brain injury, psychological symptoms, and suicide risk in a clinical sample of deployed military personnel. JAMA psychiatry 70, 686–691 (2013). 10.1001/jamapsychiatry.2013.1093

5 Hoge, C. W. et al. Mild traumatic brain injury in U.S. Soldiers returning from Iraq. The New England journal of medicine 358, 453–463 (2008).

6 Taylor, B. C. et al. Prevalence and costs of co-occurring traumatic brain injury with and without psychiatric disturbance and pain among Afghanistan and Iraq War Veteran V.A. users. Medical care 50, 342–346 (2012). 10.1097/MLR.0b013e318245a558

7 Dikmen, S. S., Bombardier, C. H., Machamer, J. E., Fann, J. R. & Temkin, N. R. Natural history of depression in traumatic brain injury. Archives of physical medicine and rehabilitation 85, 1457–1464 (2004).

8 Griffin, B. J. et al. Moral Injury: An Integrative Review. Journal of traumatic stress 32, 350–362 (2019). 10.1002/jts.22362

9 Hoge, C. W. & Castro, C. A. Treatment of generalized war-related health concerns: placing TBI and PTSD in context. JAMA : the journal of the American Medical Association 312, 1685–1686 (2014). 10.1001/jama.2014.6670

10 Pietrzak, R. H., Johnson, D. C., Goldstein, M. B., Malley, J. C. & Southwick, S. M. Perceived stigma and barriers to mental health care utilization among OEF-OIF veterans. Psychiatr Serv 60, 1118–1122 (2009). 10.1176/ps.2009.60.8.1118

11 Mac Donald, C. L., et al. Early Clinical Predictors of 5-Year Outcome After Concussive Blast Traumatic Brain Injury. JAMA neurology 74, 821–829 (2017). 10.1001/jamaneurol.2017.0143

12 Seal, K. H., Bertenthal, D., Miner, C. R., Sen, S. & Marmar, C. Bringing the war back home: mental health disorders among 103,788 US veterans returning from Iraq and Afghanistan seen at Department of Veterans Affairs facilities. Archives of internal medicine 167, 476–482 (2007). 10.1001/archinte.167.5.476

13 Tanielian, T. L. & Jaycox, L. H. Invisible Wounds of War: Psychological and Cognitive Injuries, Their Consequences, and Services to Assist Recovery. (RAND Corporation 2008).

14 Butler, A. C., Chapman, J. E., Forman, E. M. & Beck, A. T. The empirical status of cognitive-behavioral therapy: a review of meta-analyses. Clinical psychology review 26, 17–31 (2006). 10.1016/j.cpr.2005.07.003

15 Cuijpers, P., Cristea, I. A., Karyotaki, E., Reijnders, M. & Huibers, M. J. How effective are cognitive behavior therapies for major depression and anxiety disorders? A meta-analytic update of the evidence. World Psychiatry 15, 245–258 (2016). 10.1002/wps.20346

16 Elkin, I. et al. National Institute of Mental Health Treatment of Depression Collaborative Research Program. General effectiveness of treatments. Archives of general psychiatry 46, 971–982; discussion 983 (1989). 10.1001/archpsyc.1989.01810110013002

17 Shapiro, D. A. et al. Effects of treatment duration and severity of depression on the effectiveness of cognitive-behavioral and psychodynamic-interpersonal psychotherapy. Journal of consulting and clinical psychology 62, 522–534 (1994). 10.1037/0022-006x.62.3.522

18 Simon, S. S., Cordas, T. A. & Bottino, C. M. Cognitive Behavioral Therapies in older adults with depression and cognitive deficits: a systematic review. International journal of geriatric psychiatry 30, 223–233 (2015). 10.1002/gps.4239

19 Clarke, A. M., Kuosmanen, T. & Barry, M. M. A systematic review of online youth mental health promotion and prevention interventions. J Youth Adolesc 44, 90–113 (2015). 10.1007/s10964-014-0165-0

20 Gloaguen, V., Cottraux, J., Cucherat, M. & Blackburn, I. M. A meta-analysis of the effects of cognitive therapy in depressed patients. Journal of affective disorders 49, 59–72 (1998). 10.1016/s0165-0327(97)00199-7

21 Erdman, H. P., Klein, M. H. & Greist, J. H. Direct patient computer interviewing. Journal of consulting and clinical psychology 53, 760–773 (1985). 10.1037//0022-006x.53.6.760

22 Selmi, P. M., Klein, M. H., Greist, J. H., Sorrell, S. P. & Erdman, H. P. Computer-administered cognitive-behavioral therapy for depression. The American journal of psychiatry 147, 51–56 (1990). 10.1176/ajp.147.1.51

23 Arnberg, F. K., Linton, S. J., Hultcrantz, M., Heintz, E. & Jonsson, U. Internet-delivered psychological treatments for mood and anxiety disorders: a systematic review of their efficacy, safety, and cost-effectiveness. PloS one 9, e98118 (2014). 10.1371/journal.pone.0098118

24 Christensen, H. & Griffiths, K. M. The prevention of depression using the Internet. The Medical journal of Australia 177, S122–125 (2002). 10.5694/j.1326-5377.2002.tb04871.x

25 Christensen, H., Griffiths, K. M. & Korten, A. Web-based cognitive behavior therapy: analysis of site usage and changes in depression and anxiety scores. J Med Internet Res 4, e3 (2002). 10.2196/jmir.4.1.e3

26 Karyotaki, E. et al. Efficacy of Self-guided Internet-Based Cognitive Behavioral Therapy in the Treatment of Depressive Symptoms: A Meta-analysis of Individual Participant Data. JAMA psychiatry 74, 351–359 (2017). 10.1001/jamapsychiatry.2017.0044

27 Kelders, S. M., Bohlmeijer, E. T., Pots, W. T. & van Gemert-Pijnen, J. E. Comparing human and automated support for depression: Fractional factorial randomized controlled trial. Behaviour research and therapy 72, 72–80 (2015). 10.1016/j.brat.2015.06.014

28 Rathbone, A. L., Clarry, L. & Prescott, J. Assessing the Efficacy of Mobile Health Apps Using the Basic Principles of Cognitive Behavioral Therapy: Systematic Review. J Med Internet Res 19, e399 (2017). 10.2196/jmir.8598

29 Firth, J. et al. The efficacy of smartphone-based mental health interventions for depressive symptoms: a meta-analysis of randomized controlled trials. World Psychiatry 16, 287–298 (2017). 10.1002/wps.20472

30 de Graaf, L. E., Huibers, M. J., Riper, H., Gerhards, S. A. & Arntz, A. Use and acceptability of unsupported online computerized cognitive behavioral therapy for depression and associations with clinical outcome. Journal of affective disorders 116, 227–231 (2009). 10.1016/j.jad.2008.12.009

31 Al Sayegh, A., Sandford, D. & Carson, A. J. Psychological approaches to treatment of postconcussion syndrome: a systematic review. Journal of neurology, neurosurgery, and psychiatry 81, 1128–1134 (2010). 10.1136/jnnp.2008.170092

32 Fann, J. R., Hart, T. & Schomer, K. G. Treatment for depression after traumatic brain injury: a systematic review. Journal of neurotrauma 26, 2383–2402 (2009). 10.1089/neu.2009.1091

33 Kwan, V., Bihelek, N., Anderson, V. & Yeates, K. A Review of Smartphone Applications for Persons With Traumatic Brain Injury: What Is Available and What Is the Evidence? The Journal of head trauma rehabilitation 34, E45–E51 (2019). 10.1097/HTR.0000000000000425

34 Wenzel, A., Brown, G. K., & Karlin, B. E. Cognitive Behavioral Therapy for Depression in Veterans and Military Servicemembers: Therapist Manual. . (Department of Veterans Affairs, 2011).

35 Edmondson, E., Fann, J. R., Ludman, E., Dyer, J., & Bombardier, C. H. CBT-TBI Counseling Program: A Clinical Treatment Approach from Project LIFT, Therapist Manual., (University of Washington., 2014).

36 Fann, J. R., & Bombardier, C. H.. CBT-TBI Counseling Program: A Clinical Treatment Approach from Project LIFT, Patient Manual. . (University of Washington, 2014).

37 Simon, G. E., Ludman, E. J., Tutty, S., Operskalski, B. & Von Korff, M. Telephone psychotherapy and telephone care management for primary care patients starting antidepressant treatment: a randomized controlled trial. JAMA : the journal of the American Medical Association 292, 935–942 (2004). 10.1001/jama.292.8.935

38 Ritterband, L. M. et al. Efficacy of an Internet-based behavioral intervention for adults with insomnia. Archives of general psychiatry 66, 692–698 (2009). 10.1001/archgenpsychiatry.2009.66

39 Thorndike, F. P. et al. A randomized controlled trial of an internet intervention for adults with insomnia: effects on comorbid psychological and fatigue symptoms. Journal of clinical psychology 69, 1078–1093 (2013). 10.1002/jclp.22032

40 Christensen, H. et al. Effectiveness of an online insomnia program (SHUTi) for prevention of depressive episodes (the GoodNight Study): a randomised controlled trial. The lancet. Psychiatry 3, 333–341 (2016). 10.1016/S2215-0366(15)00536-2

41 Wenzel, A., Brown, G. K., & Karlin, B. E. Cognitive Behavioral Therapy for Depression in Veterans and Military Servicemembers: Therapist Manual. (U.S. Department of Veterans Affairs., 2011).

42 Fitzpatrick, K. K., Darcy, A. & Vierhile, M. Delivering Cognitive Behavior Therapy to Young Adults With Symptoms of Depression and Anxiety Using a Fully Automated Conversational Agent (Woebot): A Randomized Controlled Trial. JMIR Ment Health 4, e19 (2017). 10.2196/mental.7785

43 Kroenke, K., Spitzer, R. L. & Williams, J. B. The PHQ-9: validity of a brief depression severity measure. Journal of general internal medicine 16, 606–613 (2001). 10.1046/j.1525-1497.2001.016009606.x

44 Prescott, M. R. et al. Validation of lay-administered mental health assessments in a large Army National Guard cohort. International journal of methods in psychiatric research 23, 109–119 (2014). 10.1002/mpr.1416

45 Mackinnon, A., Griffiths, K. M. & Christensen, H. Comparative randomised trial of online cognitive-behavioural therapy and an information website for depression: 12-month outcomes. The British journal of psychiatry : the journal of mental science 192, 130–134 (2008). 10.1192/bjp.bp.106.032078

46 Lowe, B., Kroenke, K., Herzog, W. & Grafe, K. Measuring depression outcome with a brief self-report instrument: sensitivity to change of the Patient Health Questionnaire (PHQ-9). Journal of affective disorders 81, 61–66 (2004). 10.1016/S0165-0327(03)00198-8

47 Fine, T. H. et al. Validation of the telephone-administered PHQ-9 against the in-person administered SCID-I major depression module. Journal of affective disorders 150, 1001–1007 (2013). 10.1016/j.jad.2013.05.029

48 Erbe, D., Eichert, H. C., Rietz, C. & Ebert, D. Interformat reliability of the patient health questionnaire: Validation of the computerized version of the PHQ-9. Internet Interv 5, 1–4 (2016). 10.1016/j.invent.2016.06.006

49 Tulsky, D. S. et al. TBI-QOL: Development and Calibration of Item Banks to Measure Patient Reported Outcomes Following Traumatic Brain Injury. The Journal of head trauma rehabilitation 31, 40–51 (2016). 10.1097/HTR.0000000000000131

50 Blevins, C. A., Weathers, F. W., Davis, M. T., Witte, T. K. & Domino, J. L. The Posttraumatic Stress Disorder Checklist for DSM-5 (PCL-5): Development and Initial Psychometric Evaluation. Journal of traumatic stress 28, 489–498 (2015). 10.1002/jts.22059

51 Ashbaugh, A. R., Houle-Johnson, S., Herbert, C., El-Hage, W. & Brunet, A. Psychometric Validation of the English and French Versions of the Posttraumatic Stress Disorder Checklist for DSM-5 (PCL-5). PloS one 11, e0161645 (2016). 10.1371/journal.pone.0161645

52 Bastien, C. H., Vallieres, A. & Morin, C. M. Validation of the Insomnia Severity Index as an outcome measure for insomnia research. Sleep medicine 2, 297–307 (2001). 10.1016/s1389-9457(00)00065-4

53 Morin, C. M., Belleville, G., Belanger, L. & Ivers, H. The Insomnia Severity Index: psychometric indicators to detect insomnia cases and evaluate treatment response. Sleep 34, 601–608 (2011). 10.1093/sleep/34.5.601

54 Jenkins, M. M. et al. Prevalence and Mental Health Correlates of Insomnia in First-Encounter Veterans with and without Military Sexual Trauma. Sleep 38, 1547–1554 (2015). 10.5665/sleep.5044

55 Gagnon, C., Belanger, L., Ivers, H. & Morin, C. M. Validation of the Insomnia Severity Index in primary care. J Am Board Fam Med 26, 701–710 (2013). 10.3122/jabfm.2013.06.130064

56 Ashton, M. C. & Lee, K. How Well Do Big Five Measures Capture HEXACO Scale Variance? Journal of personality assessment 101, 567–573 (2019). 10.1080/00223891.2018.1448986

57 Devilly, G. J. & Borkovec, T. D. Psychometric properties of the credibility/expectancy questionnaire. Journal of behavior therapy and experimental psychiatry 31, 73–86 (2000). 10.1016/s0005-7916(00)00012-4

58 Stoyanov, S. R., Hides, L., Kavanagh, D. J. & Wilson, H. Development and Validation of the User Version of the Mobile Application Rating Scale (uMARS). JMIR Mhealth Uhealth 4, e72 (2016). 10.2196/mhealth.5849

59 Berry, K., Salter, A., Morris, R., James, S. & Bucci, S. Assessing Therapeutic Alliance in the Context of mHealth Interventions for Mental Health Problems: Development of the Mobile Agnew Relationship Measure (mARM) Questionnaire. J Med Internet Res 20, e90 (2018). 10.2196/jmir.8252

60 Malarkey, M. E. et al. Internet-Guided Cognitive Behavioral Therapy for Insomnia Among Patients With Traumatic Brain Injury: A Randomized Clinical Trial. JAMA Netw Open 7, e2420090 (2024). 10.1001/jamanetworkopen.2024.20090

61 Miller, K. E. et al. Clinician Perceptions Related to the Use of the CBT-I Coach Mobile App. Behavioral sleep medicine 17, 481–491 (2019). 10.1080/15402002.2017.1403326

62 Koffel, E. et al. A randomized controlled pilot study of CBT-I Coach: Feasibility, acceptability, and potential impact of a mobile phone application for patients in cognitive behavioral therapy for insomnia. Health Informatics J 24, 3–13 (2018). 10.1177/1460458216656472

63 Kuhn, E. et al. CBT-I Coach: A Description and Clinician Perceptions of a Mobile App for Cognitive Behavioral Therapy for Insomnia. Journal of clinical sleep medicine : JCSM : official publication of the American Academy of Sleep Medicine 12, 597–606 (2016). 10.5664/jcsm.5700

64 Zion, S. R. et al. Effects of a Cognitive Behavioral Digital Therapeutic on Anxiety and Depression Symptoms in Patients With Cancer: A Randomized Controlled Trial. JCO Oncol Pract 18, 1179–1189 (2023). 10.1200/OP.23.00210

65 Buntrock, C. et al. Effect of a Web-Based Guided Self-help Intervention for Prevention of Major Depression in Adults With Subthreshold Depression: A Randomized Clinical Trial. JAMA : the journal of the American Medical Association 315, 1854–1863 (2016). 10.1001/jama.2016.4326

66 Thorndike, F. P. et al. Development and Perceived Utility and Impact of an Internet Intervention for Insomnia. E-journal of applied psychology : clinical and social issues 4, 32–42 (2008). 10.7790/ejap.v4i2.133

67 Twomey, C. et al. A randomized controlled trial of the computerized CBT programme, MoodGYM, for public mental health service users waiting for interventions. The British journal of clinical psychology / the British Psychological Society 53, 433–450 (2014). 10.1111/bjc.12055

68 Wallace, T., Morris, J. T., Glickstein, R., Anderson, R. K. & Gore, R. K. Implementation of a Mobile Technology-Supported Diaphragmatic Breathing Intervention in Military mTBI With PTSD. The Journal of head trauma rehabilitation 37, 152–161 (2022). 10.1097/HTR.0000000000000774

69 Gartell, R., Morris, J. & Wallace, T. Feasibility of Using a Mobile App Supported Executive Function Intervention in Military Service Members and Veterans with mTBI and Co-Occurring Psychological Conditions. International journal of environmental research and public health 20 (2023). 10.3390/ijerph20032457

70 Wade, S. L. et al. Findings from a Randomized Controlled Trial of SMART: An EHealth Intervention for Mild Traumatic Brain Injury. J Pediatr Psychol 48, 241–253 (2023). 10.1093/jpepsy/jsac086

71 Sherer, M. et al. Mood Tracker: A Randomized Controlled Trial of a Self-Monitoring Intervention for Emotional Distress After Traumatic Brain Injury. The Journal of head trauma rehabilitation 40, E13–E22 (2025). 10.1097/HTR.0000000000000945

72 Ledoux, A. A. et al. Smartphone App-Delivered Mindfulness-Based Intervention for Mild Traumatic Brain Injury in Adolescents: Protocol for a Feasibility Randomized Controlled Trial. JMIR Res Protoc 13, e57226 (2024). 10.2196/57226

73 Domhardt, M. et al. Mechanisms of Change in Digital Health Interventions for Mental Disorders in Youth: Systematic Review. J Med Internet Res 23, e29742 (2021). 10.2196/29742

74 Taub, C. J. et al. Cognitive behavioral digital therapeutic effects on distress and quality of life in patients with cancer: National randomized controlled trial. Journal of consulting and clinical psychology 92, 727–741 (2024). 10.1037/ccp0000911

75 Furman, D. J. et al. Assessing the Efficacy and Safety of a Digital Therapeutic for Symptoms of Depression in Adolescents: Protocol for a Randomized Controlled Trial. JMIR Res Protoc 12, e48740 (2023). 10.2196/48740

76 Zainal, N. H., Soh, C. P., Van Doren, N. & Benjet, C. Do the effects of internet-delivered cognitive-behavioral therapy (i-CBT) last after a year and beyond? A meta-analysis of 154 randomized controlled trials (RCTs). Clinical psychology review 114, 102518 (2024). 10.1016/j.cpr.2024.102518

