## Supplemental Information for "Novel Cognitive Behavioral Therapy App for Depression in Mild Traumatic Brain Injury: A Randomized Controlled Trial"

**Supplemental Introduction and Background Information**

CBT is founded in the cognitive theory of depression, which posits that depression comes from different ways of thinking. Specifically, CBT is based on the idea that our thoughts, behaviors, and emotions influence each other. Sometimes, these interactions can become detrimental. The goal of CBT is to reframe dysfunctional thinking patterns, and in turn, negative emotions and behaviors. CBT is a structured psychotherapy approach to learn how to think in healthier ways[1]. There are many different types of negative thoughts that can exacerbate depressive symptoms, referred to as cognitive distortions. Some examples of cognitive distortions include: (1) catastrophizing, expecting the worst at all times, (2) overgeneralization, reaching a conclusion based on 1 incident or piece of evidence, and (3) fallacy of change, expecting others will change if we pressure them enough. Once patients in CBT recognize and understand the patterns of their cognitive distortions, they can start figuring out ways to correct the thought patterns [1].

It is now possible to deliver CBT content that brings out participation and engagement in the absence of a provider via computer or smartphone to much larger populations than in the past. A cohort study of approximately 450 US military SMs with head trauma found that five-years after experiencing a mTBI there was only slight improvement in depression [2]. Technology-based CBT interventions have been found to be more effective than less structured self-help programs[3]. Generally, mobile interventions are found to be well-accepted by patients for both physical and mental health interventions [4]. However, few mobile applications are held to any standards regarding their therapeutic attributes or have been the focus or empirical research studies [5] [6]. The usability and functionality of these mobile applications vary greatly and are rarely accompanied by privacy or safety policies [7]. When empirical studies examining the efficacy of these mobile health applications have been published, the small sample sizes have made it difficult to generalize findings [8]. However, there has been enough evidence gathered to warrant larger randomized control trials. Furthermore, there were several other concerning issues regarding smartphone applications for TBI including: (1) no peer-reviewed research to support the efficacy of the 70 applications, (2) lack of references for application content, (3) inappropriate marketing to laypersons not trained to interpret the findings of tools validated for use by healthcare professionals, (4) potential biases in self-report leading to possible application misuse.

### ACDC Coach Avatars

**Premise:** A key characteristic of clinical depression is the diminution of personal agency in the client's maintenance of healthy activities and relationships [9]. Depression is established and maintained by negative automatic thoughts, behavioral inactivation, avoidance, and social isolation, which are the therapeutic foci of CBT [10,11]. CBT addresses the reciprocal interactions among thoughts, feelings, and behavioral inactivation and is efficacious for treating depression within the context of an engaged and trusted therapeutic alliance [12] traditionally provided by face-to-face interaction. More recently, the attitudinal and practical barriers to CBT encountered by service members and veterans that have dissuaded many with depressive illness from seeking treatment are being addressed by both synchronous and asynchronous internet services. While a full explication of the optimal model of Internet psychotherapy is in its early stages, our project followed a well-accepted outline of factors that research has demonstrated to be both parsimonious and effective [13], which we augmented with additional features, described below.

**Content Delivery**: Brief video Vignettes using a script that follows a manualized model of treatment for depression implemented by four professional actors as "Coaches" with character development based on the personalities and lived experiences of service members and veterans, delivering performances guided by a professional television director and camera crew. Supplementary Table 1 provides brief synopses of their primary characteristics.

Per the recommendation from our military subject matter experts, realistic vulgar language was included, including military slang that has since become part of the English civilian lexicon. While some vulgar language was included in the coach scripts, the language was written so as not detract from the overall messages and lessons provided in the videos and voiceovers. The coach characters were introduced during the 'Overview' lesson, where the participants watched 4 introductory videos, each approximately 90 seconds long, for each coach. Next, the participants were prompted to choose a coach to guide them through the rest of the program. Throughout the rest of the lessons, coach videos were included to serve as examples or explain that week's personal challenge. The participants followed a track with a particular coach and also had the opportunity to switch to other coaches as pertinent needs arose. In addition to allowing self-identification with a particular coach, use of multiple coaches provided the opportunity for the curriculum to be deepened on any topic through multiple presentations. For example, "helplessness" is an important topic in CBT-D and was addressed by all four coaches. The participants were encouraged to examine each coach's perspective and "take what you can use and leave the rest," a common communication in CBT-D group therapy. At the end of each lesson, the coaches engaged with the participants by asking them how they are feeling, and questions about their motivation. Throughout the week, daily reminder notifications from the participants' chosen coach appeared to remind them to complete that week's lesson or that day's homework.Communications between the research team and participants were entirely remote, with no in-person interactions. Potential participants communicated directly with study staff via phone, email, or text messaging to learn about the study and ask questions. Study staff conducted all activities in private rooms using secure internet connections via approved electronic devices. Participants were encouraged to schedule study activities during times when they had privacy and limited distractions. The study team recommended that participants use secure password-protected Internet connections and not public WiFi when completing study activities online.  Participants were informed that records of their participation in this research study may only be disclosed in accordance with state and federal law, including the Federal Privacy Act, 5 U.S.C.552a, and its implementing regulations. Participants were identified by a code number, the key to which was accessible only to the investigators. The information gathered during this study was and will be kept confidential to the extent that the law allows. The participants were informed that the results may be published for scientific purposes, provided their identity is not revealed. During informed consent, research staff told potential participant that they would be randomly assigned to either CBT-D or education, but did not specify the hypotheses. Baseline information was obtained electronically or via telephone interview with trained study personnel. The research staff members were blinded to randomized assignment. Randomization occurred via a computer-generated list without stratification or blocking. All participants provided written informed consent in an electronic fashion. After the study started, they could communicate with study staff for technical problems and safety concerns, but there was no therapy or education provided by the study staff. Participants received automated reminders to use their applications up to 2x twice per week, unless they requested more frequent reminders.  Safety concerns were addressed within 1 one working day by a senior clinician, either a licensed clinical psychologist or board-certified neurologist with specialization in TBI. The apps included the phone number for the Veterans Crisis Line (800-273-8255, later changed to 988)

**Supplementary Table 1. Key Characteristics of the Coaches**

| Coach | Background | Challenges | Coping Mechanisms |
|---|---|---|---|
| **Billy** | White, 50 years old, served in 1st Gulf War, divorced twice, one 16-year-old daughter, past suicide attempt | Struggles with "stinking thinking," anger, self-criticism, and suicidal thoughts. | Uses **Behavioral Activation (BA)** and focuses on "doing" to combat negative thoughts and feelings. Developed the "**Radio DJ**" technique to name and tame self-critical thoughts. |
| **Jasmine** | Black, multi-generational military family, former intel officer forced into retirement after sexual assault by XO | Experienced shame and reluctance to seek help due to past judgment of her mother's depression, self-blame, and anxiety. | Uses "**Screensaver**" to replace negative thoughts with meaningful activities and "**Negative Thought Diving**" to score her actions and combat self-blame. |
| **Edwards** | Black, 35 years old, Army medic, single father, nursing student, partner left after baby was born | Depression manifests as anger and frustration, struggles with emotional self-control and stress management. | Developed "**Aerial Recon**" and "**Treating Myself Like I Treat My People**" strategies. |
| **Alicia** | Black, Hispanic, 29 years old, Marine Corps veteran, two children, stay-at-home mom | Depression manifests as anxiety, especially related to parenting and lack of self-care. | Embraces her **assertiveness** and **humor** as two of her coping mechanisms. |

A representative HEXACO profile for each coach guided a set of design principles and criteria focused on their backgrounds, challenges, and coping mechanisms. Compared with the "Big Five" personality model [14], the HEXACO model is useful for enhancing our understanding of the honesty, humility, and self-effacing traits exhibited by Service Members and Veterans who volunteer for military service in the United States. Profiles were created to guide the trajectory of each coach's experience, which the script aimed to depict vividly. Supplementary Figure 1 presents the profile of Coach Billy.

**Supplementary Figure 1**. Billy's HEXACO Profile

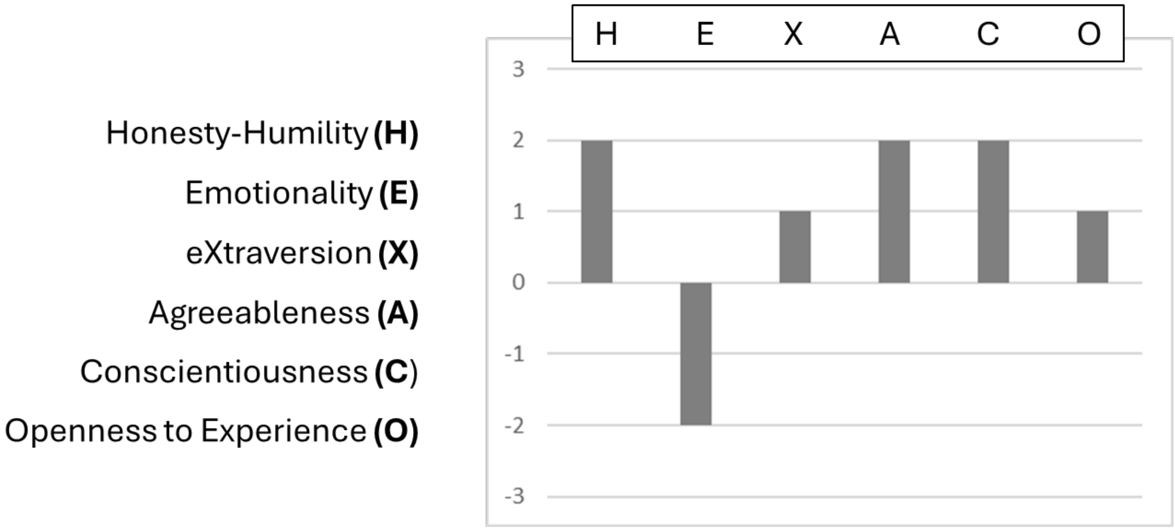


**Message Source**: The Coach profiles were crafted to provide a range of HEXACO personalities that would fit with the age, gender, and cultural background of each scripted character. We recognized that certain personality characteristics are more adaptive in "strong situations" like boot camp, advanced training, and active combat or prolonged residence in combat areas where they can become more powerfully expressed, which may be maladaptive when returning to life among civilians back home. This also guided the presentation of the CBT process with combat veterans in mind, given that American Service Members' and Veterans' concussions are often related to combat, so the warrior-civilian mismatch must be addressed in personally relevant ways. Resolution of symptoms was expected to proceed and give way to post-traumatic growth to minimize depressive disorder recidivism, evolving in a manner influenced by the personality and the lived experience of each of the coaches. Coherence with the expected personalities and cultural backgrounds of eventual recipients of the treatment program was facilitated by the active participation of a subject matter expert combat veteran, interviews with subject matter volunteers who were combat veterans, and a neurorehabilitation psychologist with extensive experience providing services to combat veterans.

**Personalization**: Early in the process, the immediate importance of coaches was to address key barriers to psychotherapy likely on the minds of curious participants early in the treatment process, given realistic pre-trauma personality profiles like those of the participants. For example, Jasmine shares an experience of feeling ashamed of her depression and reluctance to seek help, much like her adolescent experience with her mother's struggles with depression, for whom she had previously felt judgment. She describes feeling as if she had "let this happen" and was "ashamed". Her internal struggle to admit her need for help stemmed from previously being "angry with [her] mom for being depressed," leading her to feel anger with herself for experiencing the same condition. She felt that acknowledging her depression would make her "an embarrassment, just like [her] mother," further solidifying the stigma surrounding mental health care that had heretofore been an essential barrier to seeking treatment.

**Mechanisms of Change**: In addition to the manualized interventions [15,16], the coaches interacted with each other as peer counselors to use simple approximations of motivational interviewing and neurorehabilitation techniques, such as values-based goal setting and just-right challenges [17]. For example, when Billy was struggling with his goal of working out, Jasmine used motivational interviewing techniques to help him explore his values. She asked him to rate his confidence level in following through with going to the gym. As Billy explained his reasoning for his rating, he realized how important it was for him to look good in his clothes and feel like he could physically take care of himself. These were core values for Billy, and realizing this helped to motivate him to take action. In another example, Jasmine used a series of just-right challenges to rebuild her self-efficacy. The initial challenge was to write in a journal every day. This was a new activity for her and required some effort, but it was something she could accomplish. Jasmine later challenged herself to read aloud to her daughters, an activity she enjoyed before her assault but found difficult after due to problems with concentration and fatigue. Jasmine next added a weekly challenge of joining a strategic games group, gradually increasing her involvement. She reported these examples to the other coaches and modeled for participants how just-right challenges can be tailored to individual preferences and circumstances. Individuals can overcome inertia to build confidence and motivation by setting achievable goals and focusing on progress, contributing to their overall recovery and well-being. New habits such as this will likely support the long-term maintenance of emotional stability.

**Dealing with Difficult Issues:** Peer counseling occurred among the coaches. Two of the coaches, Billy and Edwards, shared their experiences with suicidal thoughts and behaviors. Billy's reflection on how suicide would sadden his daughter, Melissa, helped him commit to working on his depression. His success managing suicidal depression prompted him to become a Coach and share what he had learned with others. When Jasmine relapsed, she asked Billy to review her self-care plan, which he then helped to improve and build in self-monitoring. Edwards came into the program in deep depression with intrusive suicidal thoughts. His first steps were complicated because he was using alcohol to cope with his negative emotions and his difficult experiences. As the process unfolded, he began to address his suicidal thoughts by setting just-right challenges, including one to decrease his alcohol consumption gradually, “one day at a time,” as he shared his struggles with others by joining Alcoholics Anonymous. Both coaches emphasized that taking thoughtful action with small tasks allows them to think more clearly and helps them feel better about themselves.

**Supplementary Figure 2:** ACDC App Framework

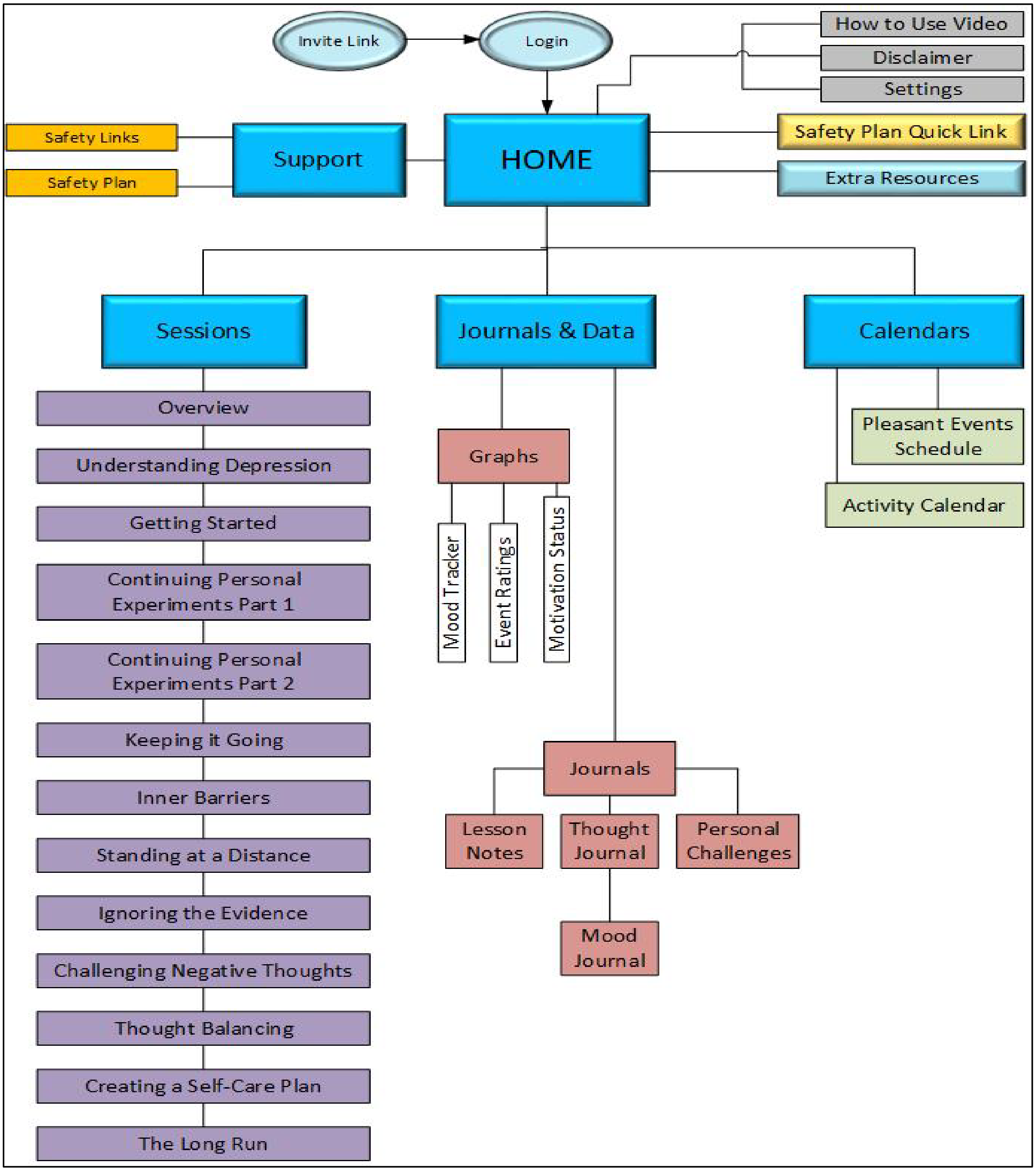

**ACDC App CBT-D Intervention lessons:**

i. **Overview.** This initial session introduced the concept of CBT to the user. It explained why and how it can be beneficial for people with concussions and depressive symptoms. Users were also introduced to the ACDC Coaches – portrayals of real-life service members and veterans who have experienced concussions and depressive symptoms and gone through CBT to help their symptoms. Users were able to watch each introduction video and choose a Coach they relate with to guide them through the intervention. The Coaches also engaged in automated motivational interviewing between lessons to ask about motivation. The Coaches' scripts were developed with the input of actual veterans, and professional actors and videographers were hired to film short segments for each lesson. At the end of the overview users completed a detailed safety plan for use throughout the program.

1. **Understanding Depression.**
    a. Lesson: What is depression?
    b. Personal Challenge: Paying closer attention to mood/mood tracking
2. **Getting Started.**
    a. Lesson: Assessing how depression affects you; learning how to conduct personal experiments
    b. Personal Challenge: Executing personal experiments and tracking emotional reactions
3. **Personal Experiments.**
    a. Lesson: Planning different personal experiments, core belief identification
    b. Personal Challenge: Personal experiments, pleasant events scheduling
4. **Continuing Your Personal Experiments.**
    a. Lesson: Internal barriers to personal experiments
    b. Personal Challenge: Personal experiments, pleasant events scheduling
5. **Adding It Up & Keeping It Going.**
    a. Lesson: Making an activity calendar
    b. Personal Challenge: Personal experiments, activity calendar
6. **Stay Active – Inner Barriers.**
    a. Lesson: How to get over internal roadblocks
    b. Personal Challenge: Pleasant events scheduling
7. **Standing at a Distance.**
    a. Lesson: Learning to watch yourself think – Negative Thoughts
    b. Personal Challenge: Thought journal, negative events
8. **Negative Focus - Ignoring the Positivity.**
    a. Lesson: Common signs of negative thinking – Assumptions
    b. Personal Challenge: Thought journal, positive events
9. **Challenging Negative Thoughts.**
    a. Lesson: Ways to challenge negative thoughts – Thought Balancing
    b. Personal Challenge: Thought journal, daily review
10. **Practicing Thought Balancing.**
    a. Lesson: Review strategies to balance thoughts, core belief modification
    b. Personal Challenge: Continue thought journal and pleasant event scheduling
11. **Self-Care.**
    a. Lesson: How to create a self-care plan
    b. Personal Challenge: Regularly review self-care plan, continue thought journal and pleasant event scheduling
12. **The Long Run.**
    a. Lesson: Review self-care plan & strategies that work best for you
    b. Personal Challenge: Continue to use the self-care plan, thought journal, and pleasant event scheduling as long as you need

**ACDC Coach Design Principles and Criteria**

**Introduction -** Although personality is generally regarded as a set of characteristics that are stable across time, research demonstrates that "strong situations" affect their development and expression. Examples of strong situations include Boot Camp and advanced training, combat patrols and active combat, and prolonged residence in combat areas. Certain personality characteristics are much more adaptive in these situations than they are elsewhere, so they become better developed and more powerfully expressed. Some of these personality characteristics are maladaptive when returning to life among civilians. For many reasons, including the mismatch between the warrior-emphasized character traits within the civilian culture back home, this can lead to a higher likelihood of depressive illness.

**Personal Relevance** - With combat veterans, CBT has efficacy for both the resolution of depressive illness symptoms and development of emotional resilience and posttraumatic growth when it addresses the warrior-civilian mismatch to facilitate transition in *personally-relevant* ways. The goal was to keep in mind “personally relevant” as scripts for the coaches were developed; this was a major challenge that in-person CBT does not face. With in-person CBT a good therapist crafts each session to keep it personally-relevant.

**Resolution of Symptoms and Posttraumatic Growth** - Successful CBT is foundational for posttraumatic growth (PTG) for several reasons, including minimizing *depressive disorder recidivism*. The goal is to set the stage for PTG during CBT by emphasizing *personal agency*; the individual is in charge of improvement with symptoms as well as with emotional growth. Although the initial goal is to pay attention to, and make progress with depressive symptoms, as symptoms wane the goal shifts maintaining compliance when participants develop values-based goals. These are distinguished from therapy-based goals that focus on resolution of symptoms. By focusing on values-based goals, the participant can integrate resolution of symptoms. Conversely, if the focus is only therapy-based goals the participant makes progress up to the point at which symptoms have become managed. Experienced therapists have a term for this: “Patients don’t want to get better, they just want to feel better.” Resolution of depressive symptoms is crucial, but because many participants developed depressive illness as a function of their goal-oriented success in combat, it will be important to help them become focused on goal-oriented success in civilian life.

**Advantage of Coach over Expert Therapist** - Coaches can help the participant better relate to the CBT experiences, thereby improving compliance. The coaches’ personality profiles must be more amenable to posttraumatic growth than the participant is currently experiencing, while at the same time having the coach demonstrate respect for the participant’s current self- formulation. The goal is to use modeling behavior offered by the coach in which the coach reflects on his or her less-adaptive self-formulation and behaviors as expressions of the coach’s pre-treatment maladaptive development. However, the goal is to be absolutely clear that the coaches’ messages have been crafted based on real-world combat veterans to reduce the risk of losing credibility.

It’s important to keep in mind that the better CBT professionals act as coaches and guides and educators rather than taking the problematic role of “expert therapist.” People who fail CBT often note the difficulties that they had with the person providing CBT in a too-directive expert therapist role. CBT focuses on equipping participants to become their own therapist by developing understanding of the relationships among thinking and feeling and behaving within the context of a personal formulation that is maladaptive. The key issue for good psychotherapy is *respect for the client*, which can be quite fragile when the directive and structured nature of CBT is mishandled by the therapist.

The directive nature of CBT can be handled in the app by a combination of a *suggestive stance*, a *thorough explanation of the rationale* for each step in the CBT process, and regular *non-authoritarian encouragement and approbation* for each of the assignments.

Because the app is fundamentally structured, this has to be called out explicitly to resolve the ambivalence that participants will naturally experience. During in- person therapy, a therapist may often ask, “Does this make sense?” and “How does this fit with your experience?” and “How did that work for you?” to recognize and honor the client’s personal experience. In the app, feature is to point to differences among the coaches, with each coach having a different response to each of the major steps in the CBT process as a way to invite the participant to seek and self-customize the experience. This is presented as an option: develop a “relationship” with the primary coach, but recognize that other coaches may also be helpful.

**ACDC Coaches**

***Billy Tilden***

- White, not Hispanic, Male
- 50 years old
- OIF combat vet
- 2009 Army National Guard unit deployed to Ramadi, Iraq
- While deployed, experienced multiple TBIs due to mortar attacks and blast waves from RPGs
- Describes depression as what he calls "stinkin' thinkin', easy to blame yourself for not moving forward
- Describes talking to his daughter even when he didn't feel like it as helpful

***Lt Commander Jasmine Matthews, Retired***

- Black, Hispanic
- 47 years old
- Married mother of two
- 8th generation in her family to join the military
- 2nd generation US Naval Academy graduate
- Physically and sexually assaulted by her XO, resulting in concussion and early retirement.
- Describes depression as feeling down, sad, hopeless, along with loss of confidence or feeling worthless
- Describes feeling anxious, irritable, or angry as parts of depression
- Described putting on her favorite blazer on days when she was feeling particularly down, which made her feel more confident

***Sergeant Major Marcellus Victor Edwards, Jr***

- Black, not Hispanic
- 35 years old
- Concussion on patrol when Humvee hit IED
- 3-4 concussions while playing football in high school
- Describes depression making physical symptoms worse such fatigue, headache, back pain, trouble sleeping, trouble concentrating, digestive problems
- Describes getting an Alcoholics Anonymous Big Book in the mail and reading it a little bit each day

***Alicia Lopez Martinez***

- White, Hispanic, Female
- 29 years old
- Enlisted in the Marine Corps in 2009
- Deployed to Afghanistan in 2010 and 2011 as part of a female engagement team
- Ambushed by an attacker with a lead pipe to the head
- Humvee hit by an IED
- Describes depression as making every mistake and disappointment look huge, and every success looks small and temporary.
- Describes helping her neighbor by walking his dog

**Educational Control App Content:**

- What is a concussion?
- How does a concussion happen?
- How many people experience concussions?
- What are the symptoms of a concussion?
  - Physical Symptoms: headaches, sleep issues, sensitivity to light or noise, nausea, vomiting, dizziness, balance issues, blurred vision, fatigue
  - Cognitive Symptoms: problems paying attention, concentrating, remembering, or processing information
  - Emotional Symptoms: anxiety, irritability, depression
- How long do symptoms last?
- Does everyone experience post-concussive symptoms the same way?
- What is depression?
- What are the symptoms of depression?
- How does depression happen?
- Who experiences depression?
- Do concussions always cause depression?
- How long do symptoms last?
- Cycle of depression & concussion
- Treatment Options
  - Learning about symptoms
  - What if I'd prefer to just wait and see if things get better?
  - Counseling
  - CBT
  - Medications
  - Transcranial magnetic stimulation
  - Treatments for other symptoms
  - Lifestyle changes: diet, exercise, sleep
  - Meditation and yoga.
  - Stress reduction
- Why do people seek care?
- When to seek help

**Communications with Participants:**

Communications were entirely remote, with no in-person interactions. Potential participants communicated directly with study staff via phone, email, or text messaging to learn about the study and ask questions. Study staff conducted all activities in private rooms using secure internet connections via approved electronic devices. Participants were encouraged to schedule study activities during times when they had privacy and limited distractions. The study team recommended that participants use secure password-protected Internet connections and not public WiFi when completing study activities online. Participants were informed that records of their participation in this research study may only be disclosed in accordance with state and federal law, including the Federal Privacy Act, 5 U.S.C.552a, and its implementing regulations. Participants were identified by a code number, the key to which was accessible only to the investigators. The information gathered during this study was and will be kept confidential to the extent that the law allows. The participants were informed that the results may be published for scientific purposes, provided their identity is not revealed. During informed consent, research staff told potential participant that they would be randomly assigned to either CBT-D or education, but did not specify the hypotheses. Baseline information was obtained electronically or via telephone interview with trained study personnel. The research staff members were blinded to randomized assignment. Randomization occurred via a computer-generated list without stratification or blocking. All participants provided written informed consent in an electronic fashion. After the study started, they could communicate with study staff for technical problems and safety concerns, but there was no therapy or education provided by the study staff. Participants received automated reminders to use their applications up to twice per week, unless they requested more frequent reminders. Safety concerns were addressed within one working day by a senior clinician, either a licensed clinical psychologist or board-certified neurologist with specialization in TBI. The apps included the phone number for the Veterans Crisis Line (800-273-8255, later changed to 988)

Participants could receive compensation for participating in this study, up to $50 in the form of gift cards. Participants could receive $25 dollars for successfully completing study measures and questionnaires (approximately 80-90% completion) at the conclusion of week 12 end of intervention assessment and an additional $25 for the completion of assessments (approximately 80-90%) at the end of week 16 follow-up assessment.

| | Allocated to CBT-D (n = 59) | Allocated to Control (n = 54) |
|---|---|---|
| Self-reported Age, years, mean (SD) | 46.6 (11.7) | 47.3 (10.8) |
| Self-reported Gender, n (%) | | |
| Female | 30 (50.9%) | 27 (50.0%) |
| Other | 0 (0.0%) | 1 ( 1.9%) |
| Self-reported Race, n (%) | | |
| White | 48 (81.4%) | 43 (79.6%) |
| Black or African American | 5 ( 8.5%) | 4 ( 7.4%) |
| Asian | 3 ( 5.1%) | 1 ( 1.9%) |
| American Indian or Alaskan Native | 1 (1.7%) | 0 ( 0%) |
| Other (includes multiple races) | 2 (3.4%) | 1 ( 1.9%) |
| No Response | 0 ( 0%) | 5 ( 9.3%) |
| Self-reported Ethnicity, n (%) | | |
| Hispanic or Latino | 7 (11.9%) | 8 ( 14.8%) |
| Non-Hispanic or Latino | 51 (86.4%) | 45 (83.3%) |
| No Response | 1 (1.7%) | 1 (1.9%) |
| Self-reported Educational level, n (%) | | |
| Partial degree | 2 (3.4%) | 1 (1.9%) |
| High school degree or less | 3 (5.1%) | 4 ( 7.4%) |
| Associates degree | 10 (16.9%) | 5 (9.3%) |
| Some college or college degree | 16 (27.1%) | 18 (33.3%) |
| Graduate Degree | 28(47.5%) | 26 (48.1%) |
| Self-reported Military Status | | |
| Active Duty/Full-time | 5 (8.5%) | 8 (14.8%) |
| Civilian | 23 (39.0%) | 19 (35.2%) |
| Retired | 16 (27.1%) | 16 ( 29.6%) |
| Separated/Discharged | 10 (16.9%) | 5 ( 9.3%) |
| Profile/Limited | 2 (3.4%) | 1 ( 1.9%) |
| No Response | 3 (5.1%) | 5 ( 9.3%) |
| Self-reported Employment Status | | |
| Full-time | 24 (40.7%) | 26 (48.1%) |
| Part-time | 2 (3.4%) | 5 (9.3%) |
| Self-employed | 1 (1.7%) | 2 ( 3.7%) |
| Unpaid (retired, student, disabled) | 26 (44.1%) | 16 (29.6%) |
| Unemployed | 6 (10.2%) | 3 ( 5.6%) |
| No Response | 0 (0.0%) | 2 ( 3.7%) |
| Self-reported Military Branch | | |
| Air Force | 6 (10.2%) | 1 ( 1.9%) |
| Army | 13 (22.0%) | 16 (29.6%) |
| Marine Corps | 3 (5.1%) | 4 ( 7.4%) |
| Navy | 11 (18.6%) | 9 ( 16.7%) |
| More than 1 | 1 (1.7%) | 3 ( 5.6%) |
| No Response | 25 (42.4%) | 21 (38.9%) |
| Self-reported Rank | | |
| Enlisted | 21 (35.6%) | 20 (37.0%) |
| Officer | 12 (20.3%) | 13 (24.1%) |
| No Response | 26 (44.1%) | 21 (38.9%) |
| Self-reported Military Occupation | | |
| Combat | 18 (30.5%) | 13 (24.1%) |
| Non-Combat | 24 (40.7%) | 27 (50.0%) |
| No Response | 17 (28.8%) | 14 (25.9%) |

**Supplemental Table 2:** Demographics for participants who were randomized to the study (ITT compliant group).

| | Randomized to CBT-D, completed (n = 19) | Randomized to CBT-D, did not complete (n = 40) | Randomized to control, completed (n = 23) | Randomized to control, did not complete (n = 31) |
|---|---|---|---|---|
| Baseline PHQ-9 score, mean (SD) | 13.6 (4.0) | 12.4 (4.0) | 12.3 (5.0) | 14.9 (4.9) |
| Self-reported Age, years, mean (SD) | 48.7 (13.6) | 45.6 (10.7) | 45.7 (11.1) | 48.5 (10.6) |
| Self-reported Gender, n (%) | | | | |
| Female | 9 (47.4%) | 21 (52.5%) | 12 (52.2%) | 15 (48.4%) |
| Other | 0 (0.0%) | 0 (0.0%) | 1 ( 4.3%) | 0 (0.0%) |
| Self-reported Race, n (%) | | | | |
| White | 17 (89.5%) | 31 (77.5%) | 21 (91.3%) | 22 (71.0%) |
| Black or African American | 0 ( 0%) | 5 (12.5%) | 1 ( 4.3%) | 3 (9.7%) |
| Asian | 0 ( 0%) | 3 (7.5%) | 1 ( 4.3%) | 0 (0.0%) |
| American Indian or Alaskan Native | 1 (5.2%) | 0 (0.0%) | 0 ( 0%) | 0 (0.0%) |
| Other (includes multiple races) | 1 (5.2%) | 1 (2.5%) | 0 ( 0%) | 1 (3.2%) |
| No Response | 0 ( 0%) | 0 (0.0%) | 0 ( 0%) | 5 (16.1%) |
| Self-reported Ethnicity, n (%) | | | | |
| Hispanic or Latino | 1 (5.30%) | 6 (15.0%) | 1 ( 4.3%) | 7 (22.6%) |
| Non-Hispanic or Latino | 18 (94.7%) | 33 (82.5%) | 22 (95.7%) | 23 (74.2%) |
| No Response | 0 (0.0%) | 1 (2.5%) | 0 (0.0%) | 1 (3.2%) |
| Self-reported Educational level, n (%) | | | | |
| Partial degree | 0 (0.0%) | 2 (5.0%) | 0 (0.0%) | 1 (3.2%) |
| High school degree or less | 1 (5.3%) | 2 (5.0%) | 2 ( 8.7%) | 2 (6.5%) |
| Associates degree | 4 (21.1%) | 6 (15.0%) | 4 (17.4%) | 1 (3.2%) |
| Some college or college degree | 3 (15.8%) | 13 (32.5%) | 9 (39.1%) | 9 (29.0%) |
| Graduate Degree | 11 (57.9%) | 17 (42.5%) | 8 (34.8%) | 18 (58.1%) |
| Self-reported Military Status | | | | |
| Active Duty/Full-time | 2 (10.5%) | 3 (7.5%) | 6 (26.1%) | 2 (6.5%) |
| Civilian | 9 (47.4%) | 14 (35.0%) | 8 (34.8%) | 11 (35.5%) |
| Retired | 3 (15.8%) | 13 (32.5%) | 2 ( 8.7%) | 14 (45.2%) |
| Separated/Discharged | 4 (21.1%) | 6 (15.0%) | 3 (13.0%) | 2 (6.5%) |
| Profile/Limited | 0 (0.0%) | 2 (5.0%) | 1 ( 4.3%) | 0 (0.0%) |
| No Response | 1 (5.3%) | 2 (5.0%) | 3 (13.0%) | 2 (6.5%) |
| Self-reported Employment Status | | | | |
| Full-time | 6 (31.6%) | 18 (45.0%) | 12 (52.2%) | 14 (45.2%) |
| Part-time | 1 (5.3%) | 1 (2.5%) | 3 (13.0%) | 2 (6.5%) |
| Self-employed | 0 (0.0%) | 1 (2.5%) | 1 ( 4.3%) | 1 (3.2%) |
| Unpaid (retired, student, disabled) | 11 (57.9%) | 15 (37.5%) | 4 (17.4%) | 12 (38.7%) |
| Unemployed | 1 (5.3%) | 5 (12.5%) | 2 ( 8.7%) | 1 (3.2%) |
| No Response | 0 (0.0%) | 0 (0.0%) | 1 ( 4.3%) | 1 (3.2%) |

| | | | | |
|---|---|---|---|---|
| Self-reported Military Branch | | | | |
| Air Force | 2 (10.5%) | 3 (7.5%) | 1 ( 4.3%) | 0 (0.0%) |
| Army | 2 (10.5%) | 11 (27.5%) | 7 (30.4%) | 9 (29.0%) |
| Marine Corps | 0 (0.0%) | 3 (7.5%) | 2 ( 8.7%) | 2 (6.5%) |
| Navy | 4 (21.1%) | 7 (17.5%) | 2 ( 8.7%) | 7 (22.6%) |
| More than 1 | 1 (5.3%) | 1 (2.5%) | 2 ( 8.7%) | 1 (3.2%) |
| No Response | 10 (52.6%) | 15 (37.5%) | 9 (39.1%) | 12 (38.7%) |
| Self-reported Rank | | | | |
| Enlisted | 6 (31.6%) | 15 (37.5%) | 6 (26.1%) | 14 (45.2%) |
| Officer | 3 (15.8%) | 9 (22.5%) | 8 (34.8%) | 5 (16.1%) |
| No Response | 10 (52.6%) | 16 (40.0%) | 9 (39.1%) | 12 (38.7%) |
| Self-reported Military Occupation | | | | |
| Combat | 4 (21.1%) | 20 (50.0%) | 13 (56.5%) | 14 (45.2%) |
| Non-Combat | 8 (42.1%) | 10 (25.0%) | 3 (13.0%) | 10 (32.3%) |
| No Response | 7 (36.8%) | 10 (25.0%) | 7 (30.4%) | 7 (22.6%) |

**Supplemental Table 3**: Demographic characteristics of participants who did vs. did not complete the study interventions.

**Supplemental Figure 3:** Comparison between groups in responses to individual PHQ-9 questions**.** The analyses were conducted in a post-hoc exploratory fashion; they were not prespecified. There were no statistically significant differences between groups.

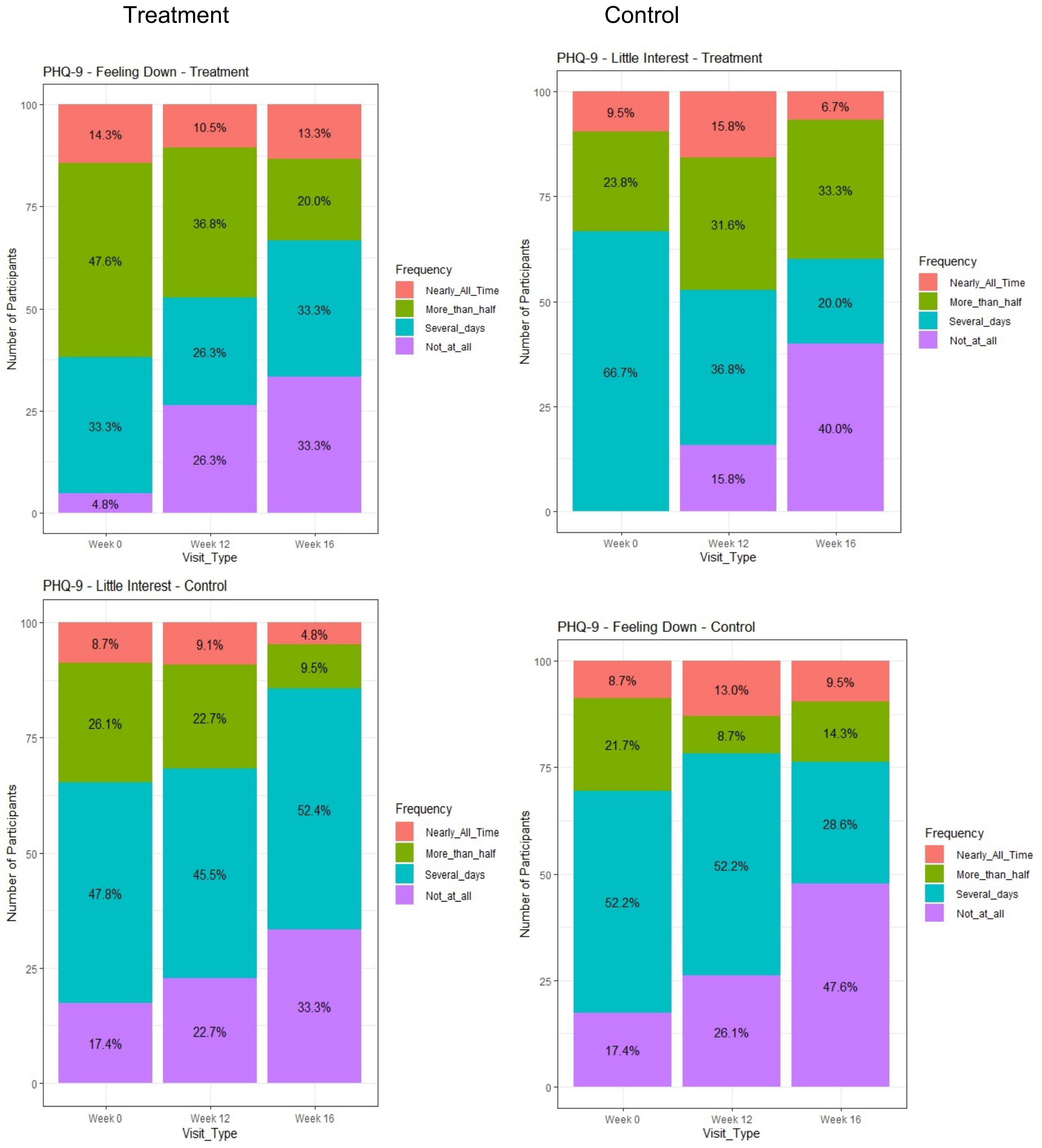

Treatment

Control

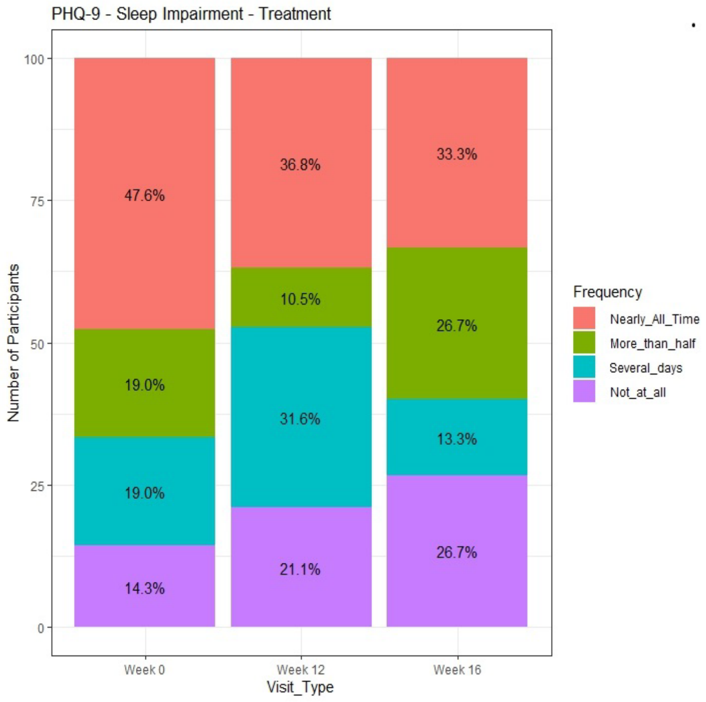


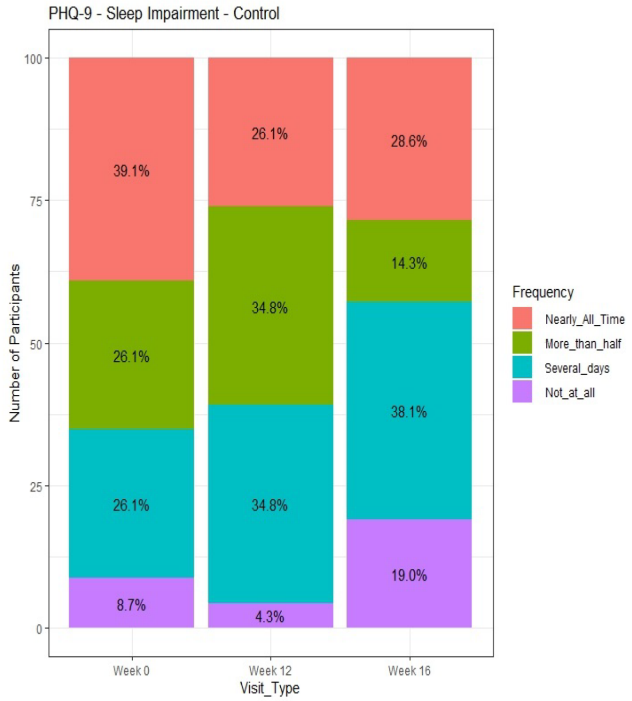


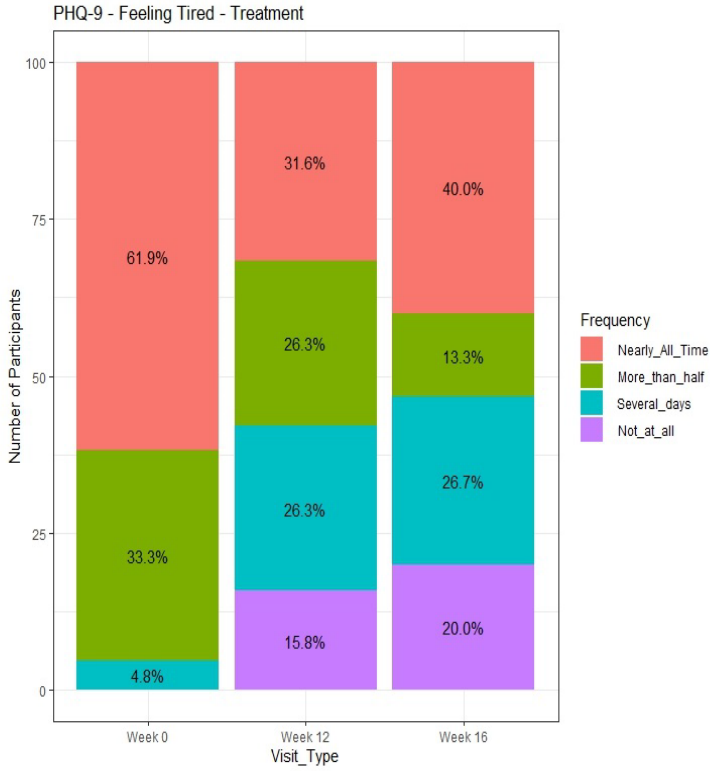


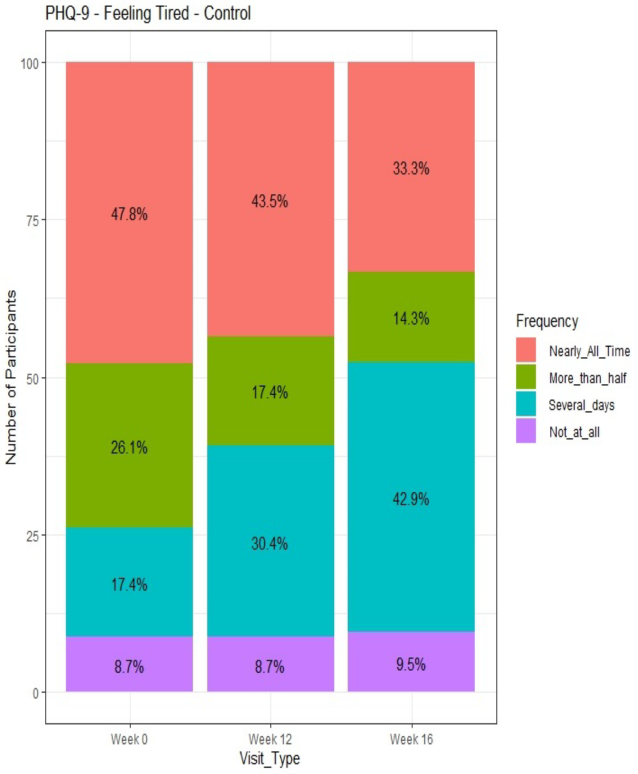

Treatment Control

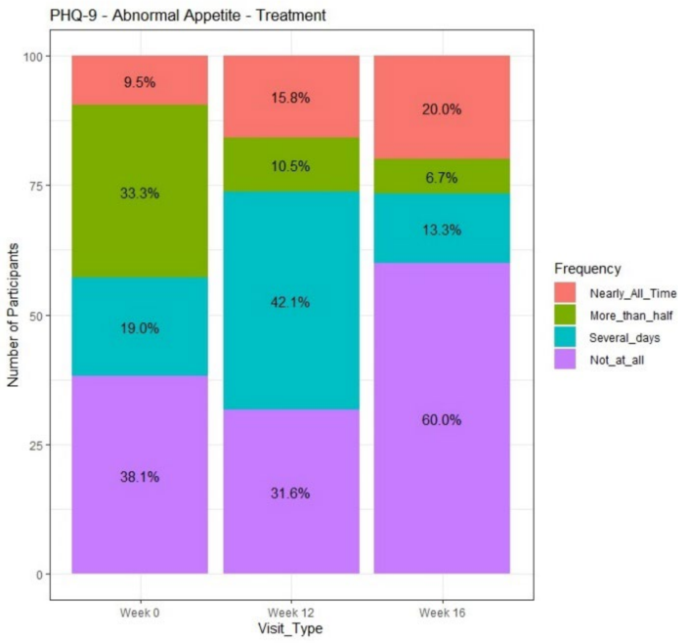


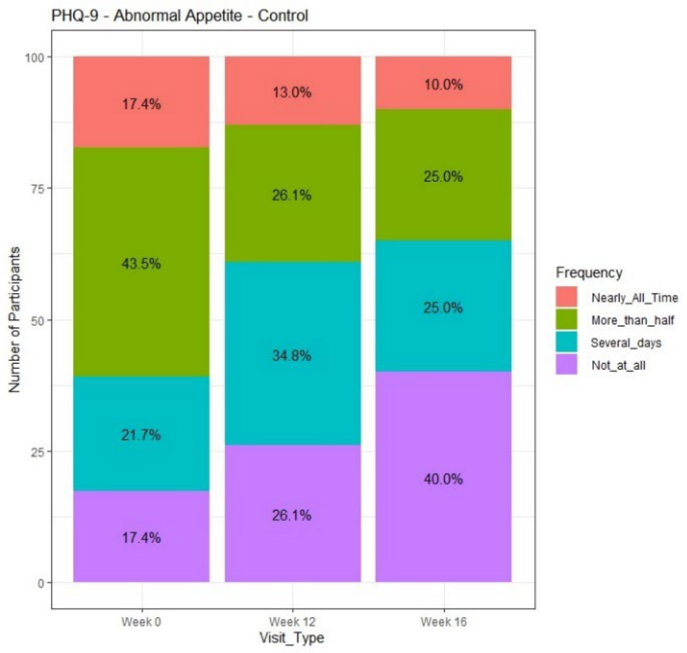


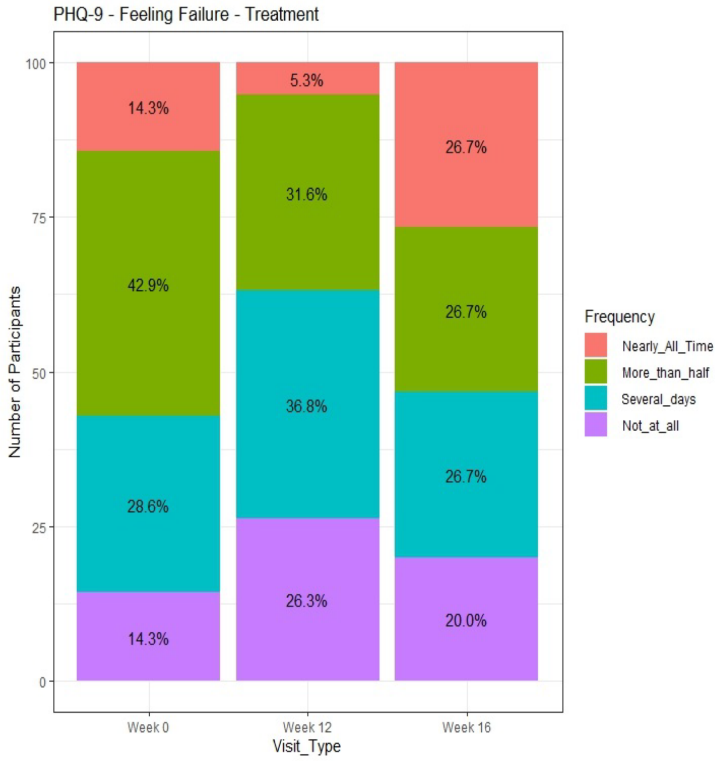


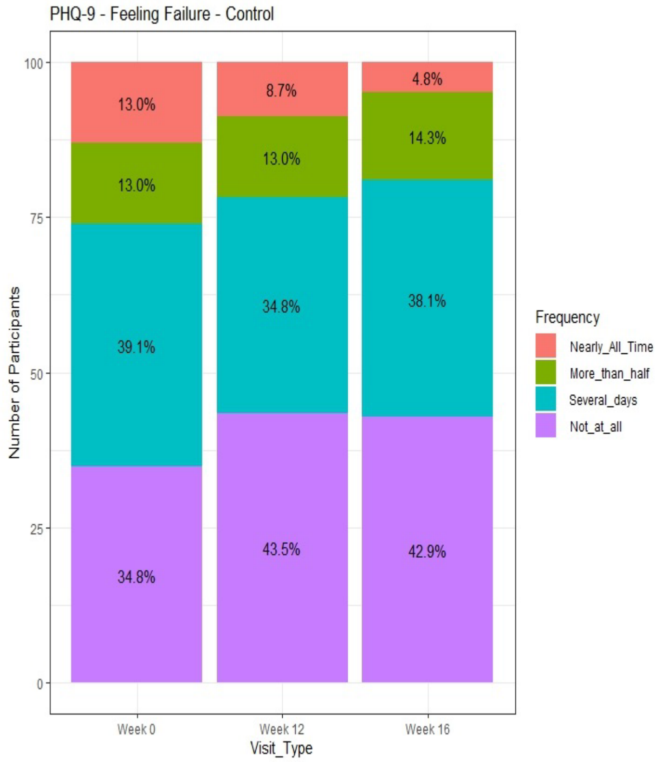

Treatment

Control

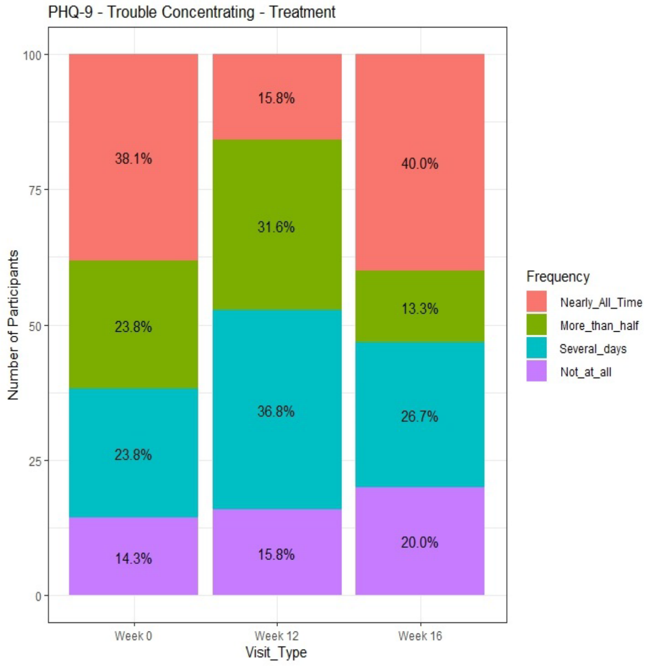


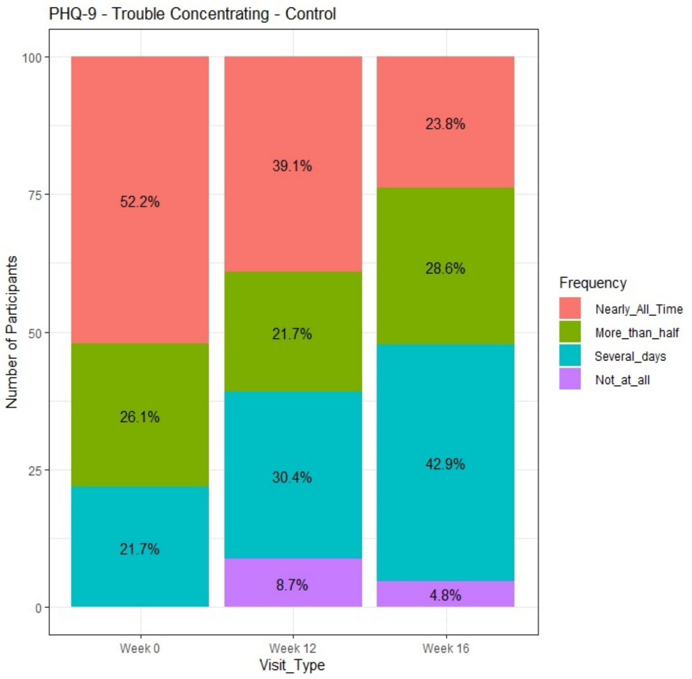


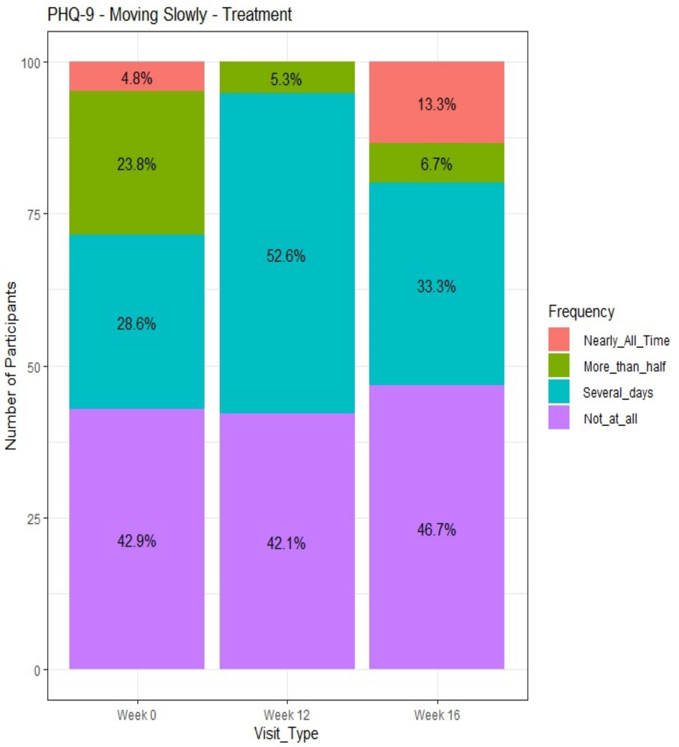


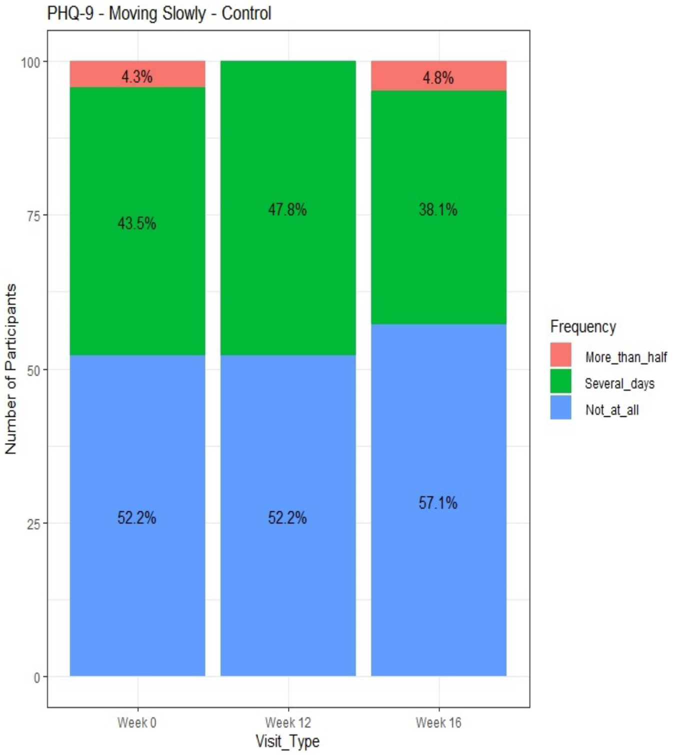

Treatment

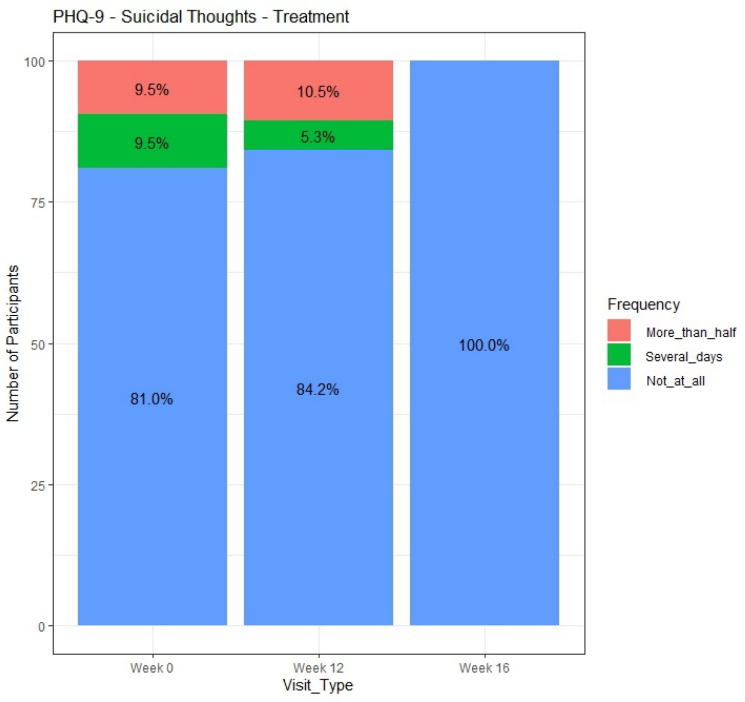


Control

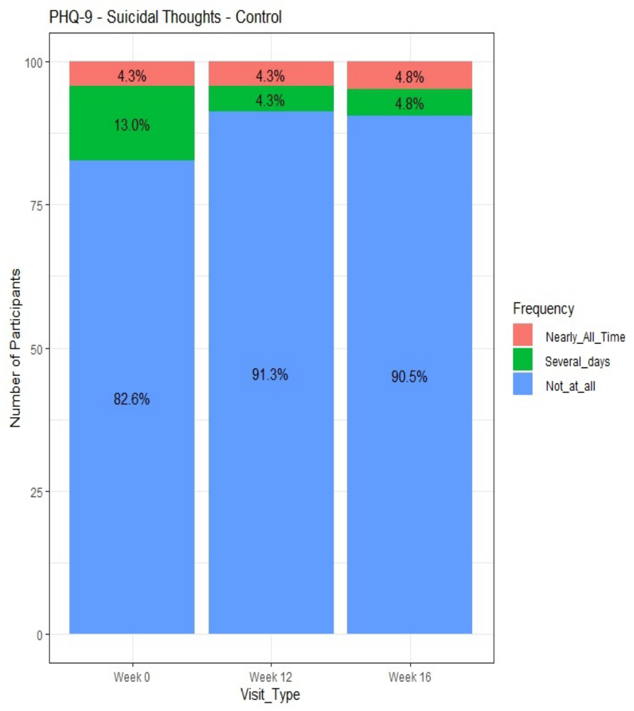

**Supplemental References**


1 Wenzel, A., Brown, G. K., & Karlin, B. E. *Cognitive Behavioral Therapy for Depression in Veterans and Military Servicemembers: Therapist Manual.* . (Department of Veterans Affairs, 2011).

2 Mac Donald, C. L. *et al.* Early Clinical Predictors of 5-Year Outcome After Concussive Blast Traumatic Brain Injury. *JAMA neurology* **74**, 821-829 (2017). https://doi.org/10.1001/jamaneurol.2017.0143

3 Titov, N. *et al.* Internet treatment for depression: a randomized controlled trial comparing clinician vs. technician assistance. *PloS one* **5**, e10939 (2010). https://doi.org/10.1371/journal.pone.0010939

4 Aguilera, A. & Muench, F. There's an App for That: Information Technology Applications for Cognitive Behavioral Practitioners. *Behav Ther (N Y N Y)* **35**, 65-73 (2012).

5 Bakker, D., Kazantzis, N., Rickwood, D. & Rickard, N. Mental Health Smartphone Apps: Review and Evidence-Based Recommendations for Future Developments. *JMIR Ment Health* **3**, e7 (2016). https://doi.org/10.2196/mental.4984

6 Tomlinson, M., Rotheram-Borus, M. J., Swartz, L. & Tsai, A. C. Scaling up mHealth: where is the evidence? *PLoS medicine* **10**, e1001382 (2013). https://doi.org/10.1371/journal.pmed.1001382

7 Huguet, A. *et al.* A Systematic Review of Cognitive Behavioral Therapy and Behavioral Activation Apps for Depression. *PloS one* **11**, e0154248 (2016). https://doi.org/10.1371/journal.pone.0154248

8 Donker, T. *et al.* Smartphones for smarter delivery of mental health programs: a systematic review. *J Med Internet Res* **15**, e247 (2013). https://doi.org/10.2196/jmir.2791

9 Bandura, A. Social cognitive theory: An agentic perspective. *Annual review of psychology* **52**, 1-26 (2001).

10 Beck, A. T. *et al. Cognitive therapy of depression*. (Guilford Publications, 2024).

11 Peterson, C. & Seligman, M. E. Causal explanations as a risk factor for depression: theory and evidence. *Psychological Review* **91**, 347 (1984).

12 Flückiger, C. *et al.* The reciprocal relationship between alliance and early treatment symptoms: A two-stage individual participant data meta-analysis. *Journal of Consulting and Clinical Psychology* **88**, 829 (2020).

13 Ritterband, L. M., Thorndike, F. P., Cox, D. J., Kovatchev, B. P. & Gonder-Frederick, L. A. A behavior change model for internet interventions. *Annals of Behavioral Medicine* **38**, 18-27 (2009).

14 McCrae, R. R. & Costa, P. T. in *Handbook of Personality: Theory and Research* Vol. John, Oliver P

Robins, Richard W

Pervin, Lawrence A Ch. 5, 159-181 (The Guilford Press, 2008).

15 Wenzel, A., Brown, G. K. & Karlin, B. E. Cognitive behavioral therapy for depression in veterans and military servicemembers: Therapist manual. *Washington, DC: US Department of Veterans Affairs* (2011).

16 Fann, J. & Bombardier, C. *CBT-TBI Counseling Program: A Clinical Treatment Approach from Project LIFT, Patient Manual*. (University of Washington, 2014).

17 Rollnick, S., Miller, W. R. & Butler, C. *Motivational interviewing in health care: Helping patients change behavior*. (Guilford Press, 2007).
